# Evolution of SARS-CoV-2 in the Rhine-Neckar/Heidelberg Region 01/2021 – 07/2023

**DOI:** 10.1101/2023.10.12.23296928

**Authors:** Christian Bundschuh, Niklas Weidner, Julian Klein, Tobias Rausch, Nayara Azevedo, Anja Telzerow, Jan-Philipp Mallm, Heeyoung Kim, Simon Steiger, Isabelle Seufert, Kathleen Börner, Katharina Bauer, Daniel Hübschmann, Katharina Laurence Jost, Sylvia Parthé, Paul Schnitzler, Michael Boutros, Karsten Rippe, Barbara Müller, Ralf Bartenschlager, Hans-Georg Kräusslich, Vladimir Benes

## Abstract

At the beginning of 2021 the monitoring of the circulating variants of SARS-CoV-2 was established in Germany in accordance with the Corona Surveillance Act (discontinued after July 2023) to allow a better containment of the pandemic, because certain amino acid exchanges (especially) in the spike protein lead to higher transmission as well as a reduced vaccination efficacy.

Therefore, our group performed whole genome sequencing applying the ARTIC protocol (currently V4) on Illumina’s NextSeq 500 platform (and starting in May 2023 on the MiSeq DX platform) for SARS-CoV-2 positive specimen from patients of the Heidelberg University Hospital (and associated hospitals) as well as the Public health office in Rhine-Neckar/Heidelberg region.

Our group sequenced a total of 26,795 SARS-CoV-2-positive samples between January 2021 and July 2023 - valid sequences, according to the requirements for sequence upload to the German electronic sequencing data hub (DESH) operated by the Robert Koch Institute (RKI), could be determined for 24,852 samples, while the lineage/clade could be identified for 25,912 samples.

While the year 2021 was very dynamic and changing regarding the circulating variants in the Rhine-Neckar/Heidelberg region with the initial non-variant of concerns, followed by A.27.RN and the rise of B.1.1.7 in winter/spring and its displacement by B.1.617.2 in spring/summer, which remained almost exclusive until the beginning of December and the first B.1.1.529 incidences, which rose to a proportion of 40 percent by the end of 2021 (and superseded B.1.617.2 by January 2022 with a proportion of over 90 percent). The years 2022 and 2023 were then dominated by B.1.1.529 and its numerous sublineages, especially BA.5 and BA.2, and more recently by the rise of recombinant variants, such as XBB.1.5. By the end of July 2023 (and since calendar week 20) the proportion of the recombinant variants amounted to 100 percent of all circulating variants in the Rhine-Neckar/Heidelberg region.

## Introduction

At the end of 2019, a novel coronavirus (2019-nCoV) - designated severe acute respiratory syndrome coronavirus 2 (SARS-CoV-2) and coronavirus disease (COVID-19) in February 2020 by the World Health Organization (WHO) - was identified as the cause of numerous pneumonia diseases in Wuhan, China (World Health Organization (WHO), 2020). Because of the rapid spread of the virus it only took a few months for the epidemic outbreak in China to evolve into a global pandemic with over 676 million confirmed COVID-19 cases by March 2023 (was discontinued afterwards) (Center for Systems Science and Engineering (Johns Hopkins University), 2023, World Health Organization (WHO), 2023).

But since only a small minority of acute COVID-19 infections were diagnosed and reported, the estimated number of COVID-19 cases might be 10 times higher according to seroprevalence studies (Clarke et al., 2022, Havers et al., 2020, Stringhini et al., 2020).

The S protein (with a size of 180–200 kDa) was identified as the most relevant protein for the entry into the host cell. It is composed of an extracellular N-terminus, a transmembrane (TM) domain (through the viral membrane) and a short intracellular C-terminus (Bosch et al., 2003). S comprises 1273 amino acids, which can be divided by the respective domains’ functions (the numbering starts from the N terminus) (Xia et al., 2020).

Since some mutations in the S protein correlated with an increased viral transmissibility, disease severity, reinfection after naturally acquired immunity, and vaccine efficacy the monitoring of circulating variants became an important epidemiological tool for the containment of the pandemic (Abdool Karim and de Oliveira, 2021).

Especially the variants of concern (VOC) showed an epidemiological advantage even early on and required the monitoring of the epidemiological distribution of the circulating SARS-CoV-2 variants. Therefore, viral genome sequencing is of crucial importance to track the emergence of mutations that define existing and new VOC.

This report shall provide the results of our group’s sequencing efforts in the Rhine-Neckar/Heidelberg region from January 2021 until the end of July 2023. We report on the local emergence of a variant designated as A.27.RN that simultaneously carried the N501Y and L452R mutations in the S gene during the first quarter of 2021, and its displacement by variants B.1.1.7., and subsequently B.1.617.2 and B.1.529 (and its numerous subvariants, especially the recombinant variants as well as BA.2.75).

## Materials and methods

### RNA purification

For SARS-CoV-2 PCR and sequencing analysis, RNA was isolated from upper respiratory tract specimens such as nasopharyngeal and oropharyngeal swabs as well as pharyngeal washes. The isolation was performed by the utilization of automated magnetic bead-based nucleic acid extraction protocols. Our group utilized either the QIASymphony, DSP Virus/Pathogen mini-Kit (Qiagen) or the Chemagic Viral DNA/RNA 300 Kit H96 (PerkinElmer) for magnetic bead RNA extraction according to the manufacturers’ protocols.

### RT-qPCR

Samples were screened for SARS-CoV-2 RNA by commercially available dual-target RT-PCR assays using automated analysis for E & orf1a/b employing Cobas 6800 (Roche Diagnostics) or analysis for E & N Gene (TIB Molbiol) or E & S gene (Altona Diagnostics) by Roche LightCycler and LightCycler II. Positive samples with a Ct value ≤35 (reduced to Ct ≤32 in week 10 because of a high rate of inconclusive results in samples with Ct>32) were subsequently characterized by whole genome sequencing. All commercial assays and devices were used according to the manufacturer’s instructions.

### SARS-CoV-2 genome sequencing

After an initial trial run until March 2021 comparing the Nextera library generation approach (Cov-seq) and the ARTIC protocol (Tyson et al., 2020), originally developed by New England Biolabs, we continued the whole genome sequencing only with the ARTIC protocol (currently V4).

The ARTIC protocol used in this study is essentially the recommended protocol, with three optimizations made during the sample upscaling using the NEBNext ARTIC SARS-CoV-2 FS Library Prep Kit for Illumina:

❖ Omission of the Magnetic Beads Clean-up step after PCR amplification of cDNA.
❖ The clean-up of adapter-ligated fragments was performed using an automated liquid-handling system.
❖ After the final PCR, the libraries were pooled and cleaned with SPRI-Beads at a 0.9x ratio (bead). All generated libraries were sequenced in pools of up to 384 samples using the Illumina NextSeq 500/MiSeq DX sequencer in paired-end mode with a read length of 75 bases.

Due to its length, only the important key points will be reported in this chapter - the complete, modified ARTIC protocol is shown in the appendix section.

The cDNA synthesis was performed by mixing the RNA sample with the LunaScript RT SuperMix and subsequently running a thermocycler program with primer annealing, synthesis, and heat inactivation steps.

The generated cDNA is amplified in two individual pool set assays (each with their respective ARTIC-SARS-CoV-2 primer mixes) in thermocyclers and then merged together.

Fragmentation (30 minutes) and end-preparation are performed in the same thermocycler run using the NEBNext Ultra II FS Enzyme Mix, resulting in fragments of approximately 100 base pairs in length. The adapter ligation is carried out using the NEBNext Adaptor for Illumina and the NEBNext Ultra II Ligation Master Mix. After an incubation phase at 20°C, (red) USER® enzyme is added, and the mixture is subsequently incubated at 37°C.

The PCR enrichment of adapter-ligated DNA is performed using the NEBNext Library PCR Master Mix and Index/Universal primers, followed by amplification in a thermocycler run.

Pooling of the libraries can be done without prior quality control, and purification is carried out using NEBNext Sample Purification Beads. The first purification step involves a 0.7-fold ratio of purification beads (based on the pool volume), followed by several incubation, ethanol wash, and drying steps. The final bead extraction is performed using TE buffer and a magnet. The second cleanup step is done with a 0.9-fold ratio of purification beads (based on the pool volume) and otherwise follows the same procedure as the first cleanup.

The last step of the library preparation is the quality control of the final pool using a Qubit DNA BR Kit and Agilent Bioanalyzer High Sensitivity DNA Reagents for sample dilution, the expected peak size is 200-250 base pairs with a 30-minute fragmentation time.

### Data analysis

#### Nextera

Sequencing data from Cov-seq were processed using the nf-core viralrecon pipeline (Ewels et al., 2020, Patel, 2020) with the following settings:

❖ NC_045512.2 genome
❖ Metagenomic protocol without primer sequence removal
❖ No duplicate filtering
❖ Minimal coverage of 20 for variant calling
❖ Maximum allele frequency of 0.9 for filtering variant calls.

Primer sequences were not removed due to the protocol’s specific amplicon design. Variant calls and reconstructed consensus sequences from iVar were used (Grubaugh et al., 2019).

#### ARTIC

For the data analysis of the ARTIC sequencing, the samples were always sequenced in paired-end mode with a read-length of 75bp using the NEBNext ARTIC library preparation kit, the detailed protocol is shown in Appendix B. For the data analysis, we applied a multi-stage data processing pipeline that included various tools for sequencing adapter trimming, alignment, quality control, mutation calling, viral genome assembly and lineage classification, which are listed in the following paragraphs.

❖ Sequencing adapter trimming

Short input fragments may cause sequencing adapters to be present in the final sequencing read. We applied trim galore with default parameters to trim off these adapters (https://zenodo.org/badge/latestdoi/62039322).

❖ Host-read contamination removal

Because of unspecific amplicon primer binding, the sequencing data may contain contaminating human derived reads. These host reads were removed using a k-mer based method, called kraken2 (Wood et al., 2019), and using alignments against the host reference (build GRCh38). We also applied kraken2 to screen for other potential lab contaminants (Wood et al., 2019).

❖ Alignment to SARS-CoV-2 reference genome

All remaining sequencing reads were aligned to the SARS-CoV-2 reference genome (NC_045512.2) using the Burrows-Wheeler Alignment tool (Li and Durbin, 2009). Alignments were sorted and indexed using SAMtools (Li et al., 2009) and quality-controlled using Alfred (Rausch et al., 2019). We masked priming regions with iVar (Grubaugh et al., 2019) to avoid a variant calling bias in such loci. Due to the overlapping amplicon design, this strategy does not cause further dropout (unobserved) regions.

❖ Variant calling

Variant calling employed FreeBayes (Garrison and Marth, 2012) and bcftools (Li, 2011) for quality-filtering and normalization of variants. All variants were annotated for their functional consequence using the Ensembl Variant Effect Predictor (McLaren et al., 2016).

❖ Consensus computation

SAMtools, bcftools, iVar and Alfred were also employed for viral consensus sequence generation and its quality control. The iVar consensus parameters and quality control stringency were based on the Robert Koch Institute (RKI) SARS-CoV-2 sequencing submission criteria, namely a 90% informative sequence, less than 5% Ns (unobserved bases), a minimum of 20x coverage and at least 90% read support for informative positions (Robert Koch Institute (RKI), 2021b).

❖ Lineage classification

Lineage and clade classification was carried out using Pangolin (Rambaut et al., 2020) and Nextclade (Hadfield et al., 2018), respectively, with default parameters. At the beginning of the project, where multiple wild-type lineages were present, we also employed a simplified typing system that only highlighted the variants of concern (at the beginning of our project: B.1.1.7, B.1.351 and P.1).

❖ Summary report generation

We applied a custom python script available in the GitHub source code repository (https://github.com/tobiasrausch/covid19) to aggregate all QC metrics, lineage labels and variants leading to amino acid changes. This report generation also entailed the computation of metadata tracking sheets and gzipped FASTA files for all viral assemblies to facilitate an immediate upload to the German electronic sequencing data hub (DESH) operated by the RKI.

In rare cases, we validated mutations by PCR and Sanger sequencing to confirm interesting sequencing results with the help of primer design and Sanger sequencing chromatogram analysis methods (Rausch et al., 2020) available at GEAR genomics.

### Quality criteria for sequencing data

Since the governmental regulation regarding the molecular genetic surveillance of SARS-CoV-2 (Coronavirus-Surveillanceverordnung – CorSurV) by Germany and Baden-Württemberg required the upload of the SARS-CoV-2 viral genome sequences to the DESH database of the RKI (Robert Koch Institute (RKI), 2021a, Robert Koch Institute (RKI), 2021b) the quality criteria required by the RKI were set as the requirement for acceptable sequencing quality of genomes analyzed with the Nextera and ARTIC protocols. The vertices of the RKI criteria for Illumina-based SARS-CoV-2 genome sequencing are presented in Table 1 (Robert Koch Institute (RKI), 2021b).

**Table 1.**
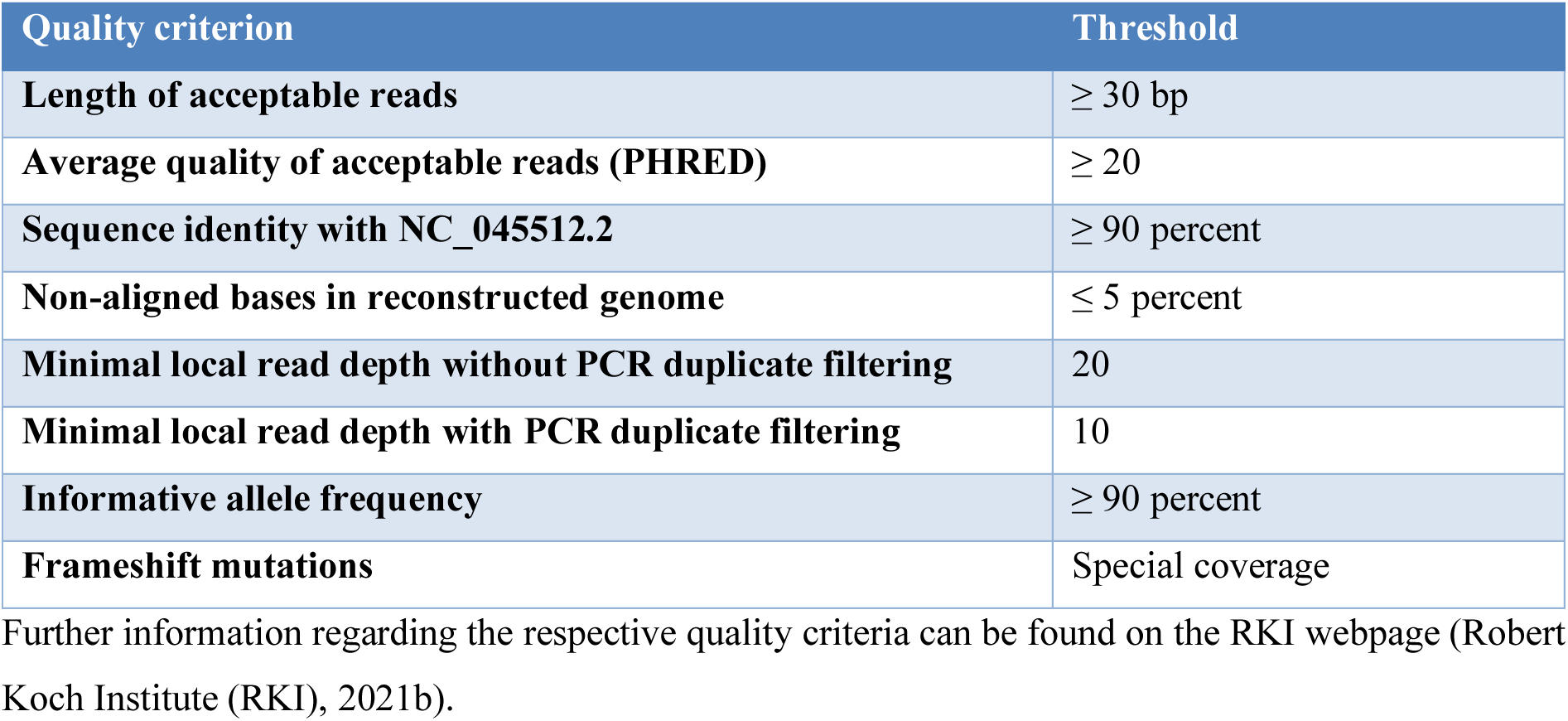
RKI quality criteria for DESH upload.

### Data availability

SARS-CoV-2 viral genome sequences have been uploaded to the DESH database (https://www.rki.de/DE/Content/InfAZ/N/Neuartiges_Coronavirus/DESH/DESH.html) of the Robert Koch Institute and will be made available via the GISAID database (Shu and McCauley, 2017).

### Ethical statement

This study was approved by the ethics committee of the Medical Faculty at the University of Heidelberg for the analysis of proband samples by whole genome sequencing of the viral RNA (S-316/2021) as well as the virus propagation and neutralization assays and use of sera from vaccinated donors (S-203/2021). Additionally, this large-scale sequencing of SARS-CoV-2 positive samples was performed in accordance with the governmental regulation regarding the molecular genetic surveillance of SARS-CoV-2 (Coronavirus-Surveillanceverordnung – CorSurV) by Germany and Baden-Württemberg.

## Results

### Method comparison Nextera – Artic

Initially, our group compared 2 different NGS protocols for SARS-CoV-2 sequencing with respect to the RKI quality criteria, the Nextera and ARTIC protocols. The respective details are provided in the material and methods section.

These comparative runs were performed with 1488 extracted RNA specimens from the department of virology of the Centre of Infectious Diseases Heidelberg and the Institute for Medical Microbiology and Hygiene of the Mannheim University Hospital from January and February 2021.

Initially, a total of 1055 specimen with a positive PCR result (and no Ct cutoff value) were analyzed for the comparative runs. As aforementioned, the required quality threshold of viral genome sequencing was set according to the RKI criteria. The respective results are presented in Table 2.

**Table 2.**
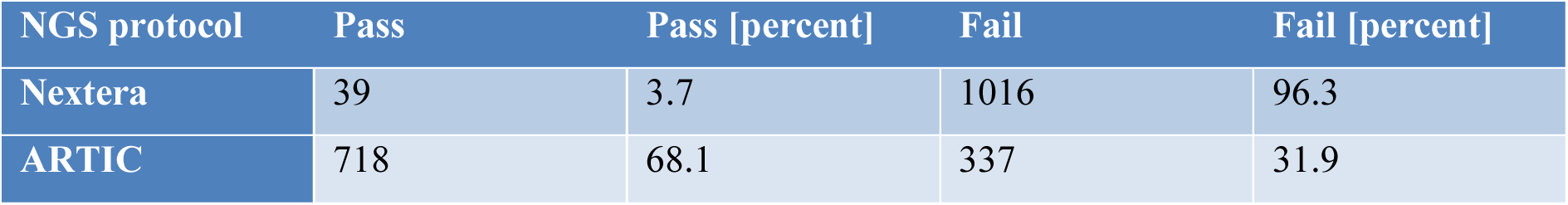
Results of the NGS runs before Ct cutoff.

| NGS protocol | Pass | Pass [percent] | Fail | Fail [percent] |
| --- | --- | --- | --- | --- |
| Nextera | 39 | 3.7 | 1016 | 96.3 |
| ARTIC | 718 | 68.1 | 337 | 31.9 |

Hence, the ARTIC protocol showed significantly better sequencing results before the establishment of a Ct cutoff value regarding the RKI criteria compared to the Nextera protocol (P-value <0.0001). Samples with Ct > 32 were excluded due to high NGS failure rates. These settings were tested with 433 specimen and the results are depicted in Table 3.

**Table 3.** Results of the NGS runs after Ct cutoff.

| NGS protocol | Pass | Pass [percent] | Fail | Fail [percent] |
| --- | --- | --- | --- | --- |
| Nextera | 197 | 45.5 | 236 | 54.5 |
| ARTIC | 396 | 91.5 | 37 | 8.5 |

After the Ct cutoff establishment, both protocols showed a significant improvement. For the Nextera protocol, the pass rate increased from 3.7 to 45.5 percent (P-value <0.0001), while the ARTIC protocol showed a pass rate of 91.5 compared to the previous 68.1 percent (P-value <0.0001).

Incorrect detection of viral genomes was only found in individual cases for both protocols.

❖ Nextera

➢ 2 mutated variants identified as wild-type
➢ 1 mutated variant identified as a different variant (B.1.1.7 → A.27.RN)
❖ ARTIC

➢ 1 mutated variant identified as wild-type

NGS results were correlated to VirSNiP melting curve assays targeting N501Y, DelHV69/70 and K417N (TIBMolbiol, Berlin, Germany) that were employed for fast detection of possible Variants of Concern.

Since the ARTIC protocol’s performance remained significantly superior to the Nextera protocol (before and) after the establishment of a Ct threshold (P-value <0.0001), all subsequent sequencing runs were performed with the ARTIC protocol on a NextSeq 500/MiSeq DX (Illumina).

### Variant distribution in 2021

Between January and May 2021, a SARS-CoV-2 variant with L452R and N501Y was found in the Rhine-Neckar region in 176 out of 3,130 specimen (peak incidence from January until March 2021 with 166 out of 1,543 samples acquired from January to March 2021). Its phylogenetic lineage (Pangolin) was A.27.RN and its first occurrence within the Rhine-Neckar/Heidelberg region was in a SARS-CoV-2 positive specimen collected during the calendar week 1 of 2021. The (initial) defining S gene mutations comprised by A.27.RN were:

❖ L18F
❖ L452R
❖ N501Y
❖ A653V
❖ H655Y
❖ D796Y and
❖ G1219V

in a D614 wild-type background. Detection of both B.1.1.7 and A.27.RN increased at a similar pace until calendar week 7, but diverged from calendar week 8 and onwards.

Since the characterization of A.27.RN was already done in a previous paper by our group (Mallm et al., 2021), A.27.RN won’t be discussed any further at this point.

Starting in calendar week 5 our group observed an increase and subsequent displacement of B.1.1.7 until Q2/2021.

The rates of B.1.351 (as well as P.3) remained constantly below the 5 percent threshold in Q1/2021. Calendar weeks 1 and 13 showed higher incidence rates with ∼10 percent for B.1.351. In Q2/2021 the rates of B.1.351 remained below the 5 percent threshold (until week 19) with only individual occurences afterwards and no more incidences after the end of Q2/2021.

While P.1 was initially thought of as the most problematic among the first three variants of concern of 2021, its incidence numbers remained low in the Rhine-Neckar/Heidelberg region and never became epidemiologically relevant - P.1 was only identified infrequently in specimens for 6 weeks before its extinction by the end of Q2/2021, similar to the incidences of B.1.351.

The presence of B.1.617.1 (Kappa, Clade 21B) was found only in a single specimen in week 14 from the Rhine-Neckar region (and in no specimen from the Mannheim region).

During Q2/2021 and within a matter of weeks, the domination by B.1.1.7 was superseded by the spread of B.1.617.2 and at the start of Q3/2021 all other variants were almost “extinct”, with only individual (resequencing) cases of long-term SARS-CoV-2-PCR-positive patients.

After its first detection in calendar week 19 the rate of B.1.617.2 significantly increased and by calendar week 26 (last week of Q2/2021) B.1.617.2 accounted for about 50% of all SARS-CoV-2 cases in the Rhine-Neckar/Heidelberg region and after another 2-4 weeks all other variants were already displaced by the B.1.617.2. Its dominance remained unchanged until the first week of December, with the detection of the first B.1.1.529 cases in specimens from returning travelers. After the initial two incidences, within a matter of only weeks (similar to the dynamics which could be observed for the displacement of B.1.1.7 and subsequently for B.1.617.2) the incidence rate rose to over 40 percent by the end of 2021.

The initial B.1.1.529 lineage was later reclassified as BA.1.

### Overview of Variant Distribution for the Year 2021

An overview of the sequenced specimen for the Rhine-Neckar region in 2021 is shown in Table 4. The case number reflects the respective variant’s sequence frequencies which fulfilled the RKI DESH criteria (Robert Koch Institute (RKI), 2021b) over the year 2021, the numbers in brackets reflect the total number of specimen which could be assigned to the respective variant following the sequencing process (including samples which passed and failed the RKI DESH criteria). The first and last discovery reflect the date of the respective variant’s first and last detection during the year 2021.

**Table 4.** SARS-CoV-2 variant statistics for the Rhine-Neckar region 2021.

| Variant | Cases | First Discovery | Last Discovery |
| --- | --- | --- | --- |
| Wild type | 722 (794) | 02/01/2021 | 26/06/2021 |
| A.27 | 170 (176) | 14/01/2021 | 19/05/2021 |
| B.1.1.7 | 1908 (1961) | 20/01/2021 | 05/08/2021 |
| including Q.1 | 43 (47) | 13/02/2021 | 11/06/2021 |
| including Q.4 | 4 (4) | 24/02/2021 | 24/02/2021 |
| B.1.351 | 51 (55) | 13/01/2021 | 08/06/2021 |
| P.1 | 11 (12) | 18/05/2021 | 25/06/2021 |
| P.3 | 5 (5) | 27/02/2021 | 20/03/2021 |
| B.1.617.1 | 2 (2) | 11/04/2021 | 11/04/2021 |
| B.1.617.2 | 4122 (4304) | 10/05/2021 | 31/12/2021 |
| including AY.4.2 | 63 (65) | 06/10/2021 | 18/12/2021 |
| B.1.1.529/BA.1 | 146 (147) | 03/12/2021 | 31/12/2021 |
| Total | 7141 (7498) |  |  |

A detailed overview of the variants classified as wild type variants (as well as their respective incidences) is shown in Appendix A.

In 2021, our group sequenced 7995 SARS-CoV-2 positive specimen from the Rhine-Neckar/Heidelberg region and as shown in Table 4 for 7498 samples the lineage and clade could be determined (93.8 percent), leaving only 497 specimens unidentifiable (6.2 percent). 7141 samples passed the minimal sequencing criteria of the RKI DESH (89.3 percent), which leaves 854 specimen that did not (10.7 percent).

The biggest (negative) gap in specimen, for which a sequence could be determined, that did not meet the RKI DESH criteria is found in the wild type group, which can be explained that the highest number of wild type samples was sequenced before the Ct value-dependent sample inclusion criterion (Ct ≤ 32) was in place and therefore more degraded RNA samples were sequenced, which resulted in overall higher failed rates regarding the RKI DESH criteria.

An overview of the number of sequenced samples per calendar week and the success rate of sequencing is depicted in Figure 1.

**Figure 1.**
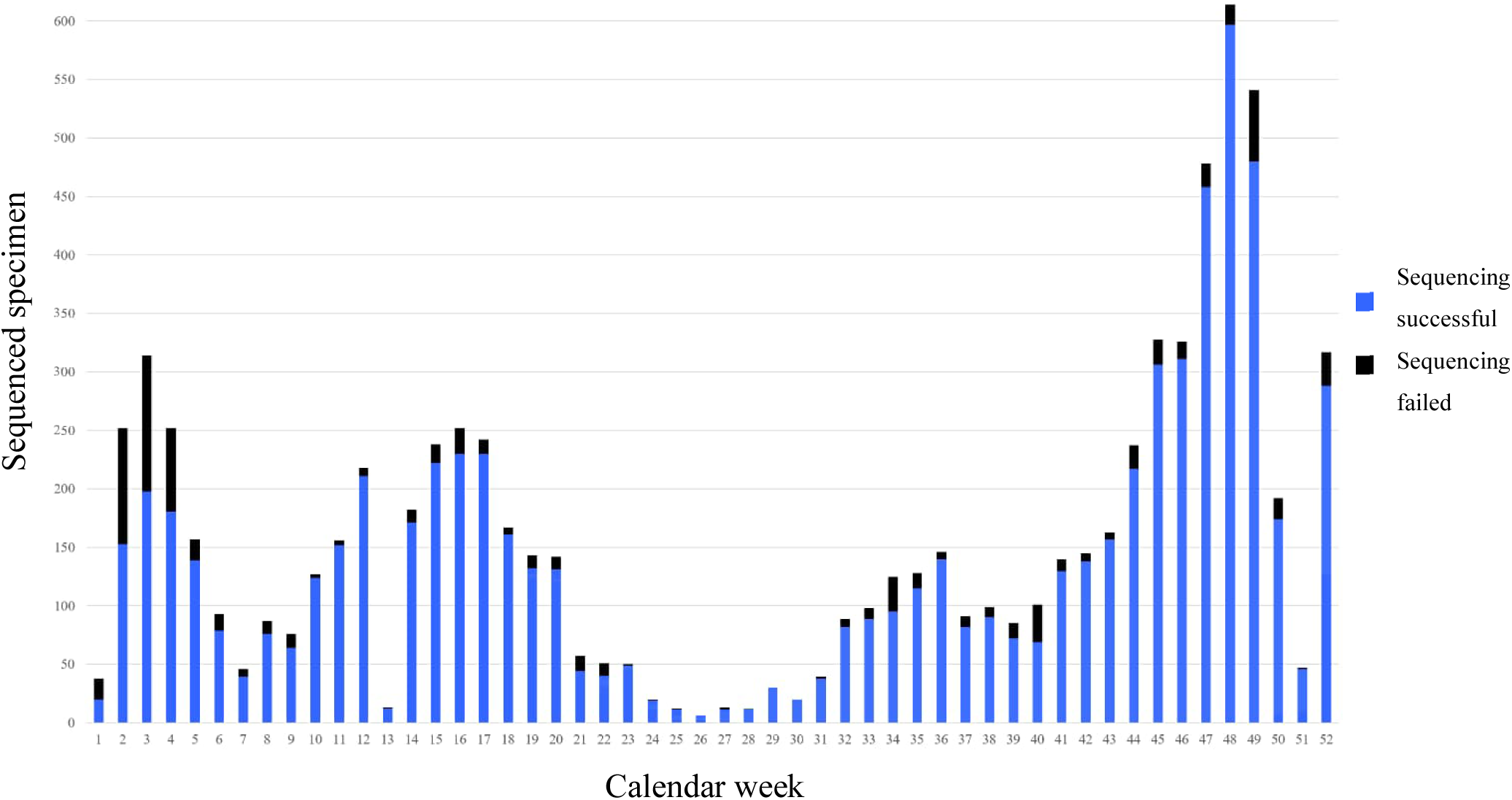
Overview of the number of successful and failed sequencings for Rhine-Neckar region specimen per calendar week for 2021.

A comprehensive overview of the variant distribution in the Rhein-Neckar/Heidelberg region for the year 2021 is presented in Figure 2.

**Figure 2.**
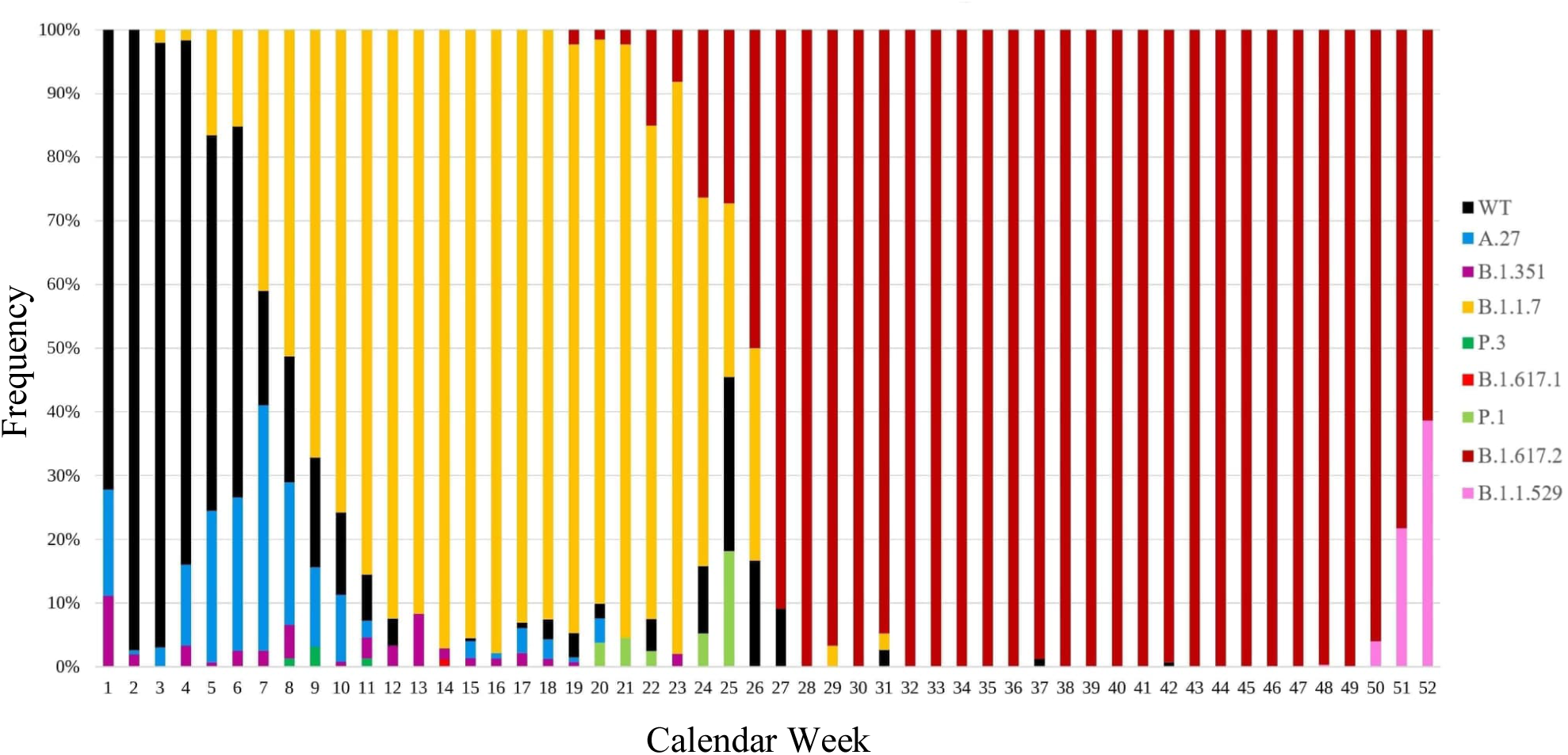
SARS-CoV-2 variant distribution per calendar week 2021.

### Overview of Variant Distribution for the Year 2022

With the beginning of the year 2022, B.1.1.529 (respectively BA.1) was almost the only relevant variant in the Rhein-Neckar/Heidelberg region and almost completely replaced B.1.617.2 by the end of January. The dominance of BA.1, however, was short-lived. Before a complete displacement of B.1.617.2 could take place, the decline of BA.1 and the rise of BA.2 began – within two months (until the end of March 2022), the prevalence of BA.1 fell below the 10% mark. However, BA.2 was also unable to compete against the subsequent variants – until the emergence of the recombinant Omicron variants, which are (currently) mainly recombinations of BA.2 sublineages.

As worldwide, BA.3 also failed to establish itself in the Rhein-Neckar/Heidelberg regions, with only isolated cases of BA.3 in calendar week 13.

Although BA.4 and BA.5 were detected almost simultaneously (between calendar weeks 15 and 19), BA.4 could not establish itself with epidemiologically relevant numbers. While the variant was not completely displaced by BA.5, it never reached a proportion of 10% of total infections per calendar week, whereas BA.5, once it surpassed the 10% mark, it took just under 4 weeks for BA.5 to account for more than 80% of total infections. For most of the second half of the year 2022, the proportion of BA.5 remained above the 90% mark. This changed only from calendar week 45 onwards, when there was a resurgence of BA.2 and the detection of recombinant Omicron variants (such as XBB).

After the first detections of BA.2.75, BQ.1, and their subvariants we saw a continuous increase in the circulating variants over the next few months. These, along with the recombinant variants of BA.2 (e.g., XBB, which emerged through recombination of BA.2.10.1 and BA.2.75), also exhibited a replication advantage over the already circulating variants. As a result, the recombinant variants became even more dominant in 2023.

An overview of the sequenced specimen for the Rhine-Neckar region in 2022 is shown in Table 5. The case number reflects the respective variant’s sequence frequencies which fulfilled the RKI DESH criteria (Robert Koch Institute (RKI), 2021b) over the year 2021, the numbers in brackets reflect the total number of specimen which could be assigned to the respective variant following the sequencing process (including samples which passed and failed the RKI DESH criteria). The first and last discovery reflect the date of the respective variant’s first and last detection during the year 2022.

**Table 5.**
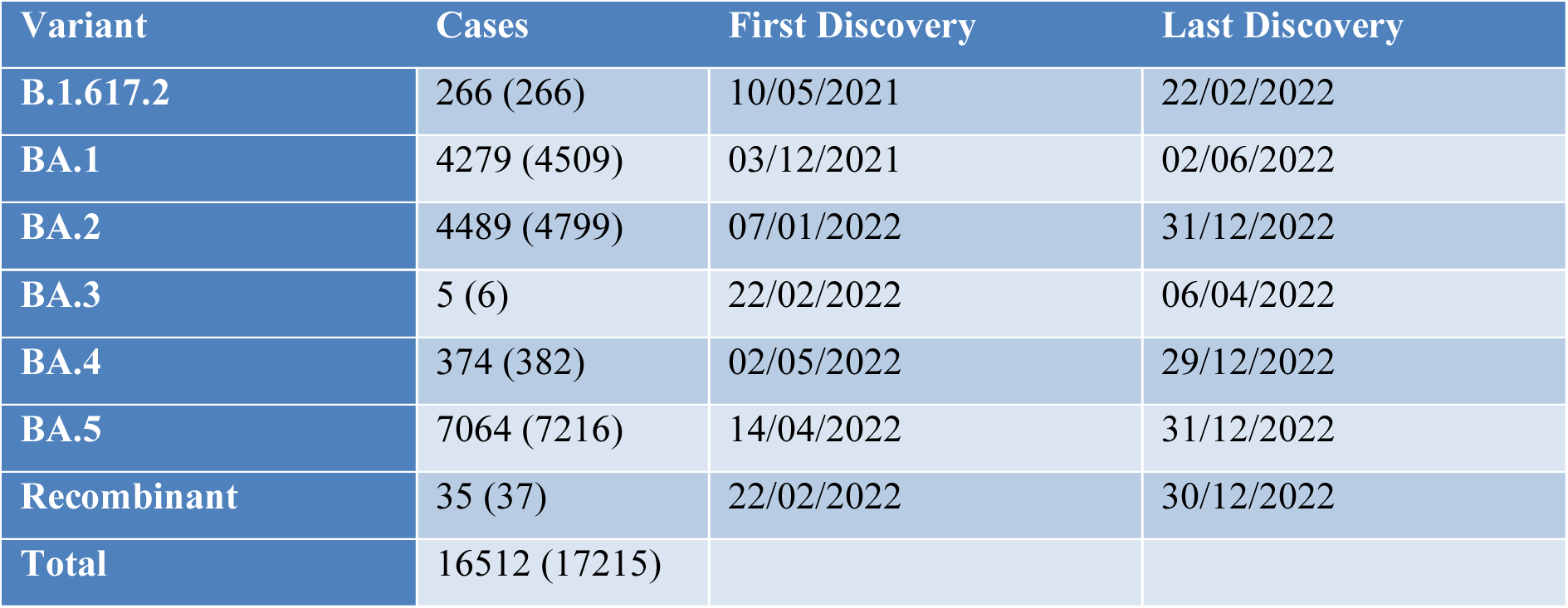
Variant Distribution in the Rhein-Neckar/Heidelberg Region in 2022.

In 2022, our group sequenced 17,455 SARS-CoV-2 positive specimen from the Rhine-Neckar/Heidelberg region and for 17,215 samples the lineage and clade were determinable (98.6 percent), leaving only 240 specimens unidentifiable (1.4 percent). 16,512 samples passed the minimal sequencing criteria of the RKI DESH (94.6 percent), which leaves 943 specimen that did not (5.4 percent).

In 2022, no significant (negative) gap was identified in sequenced samples that did not meet the RKI DESH criteria, unlike in the year 2021 (Bundschuh, 2021).

An overview of the number of sequenced samples per calendar week and the success rate of sequencing is depicted in Figure 3.

**Figure 3.**
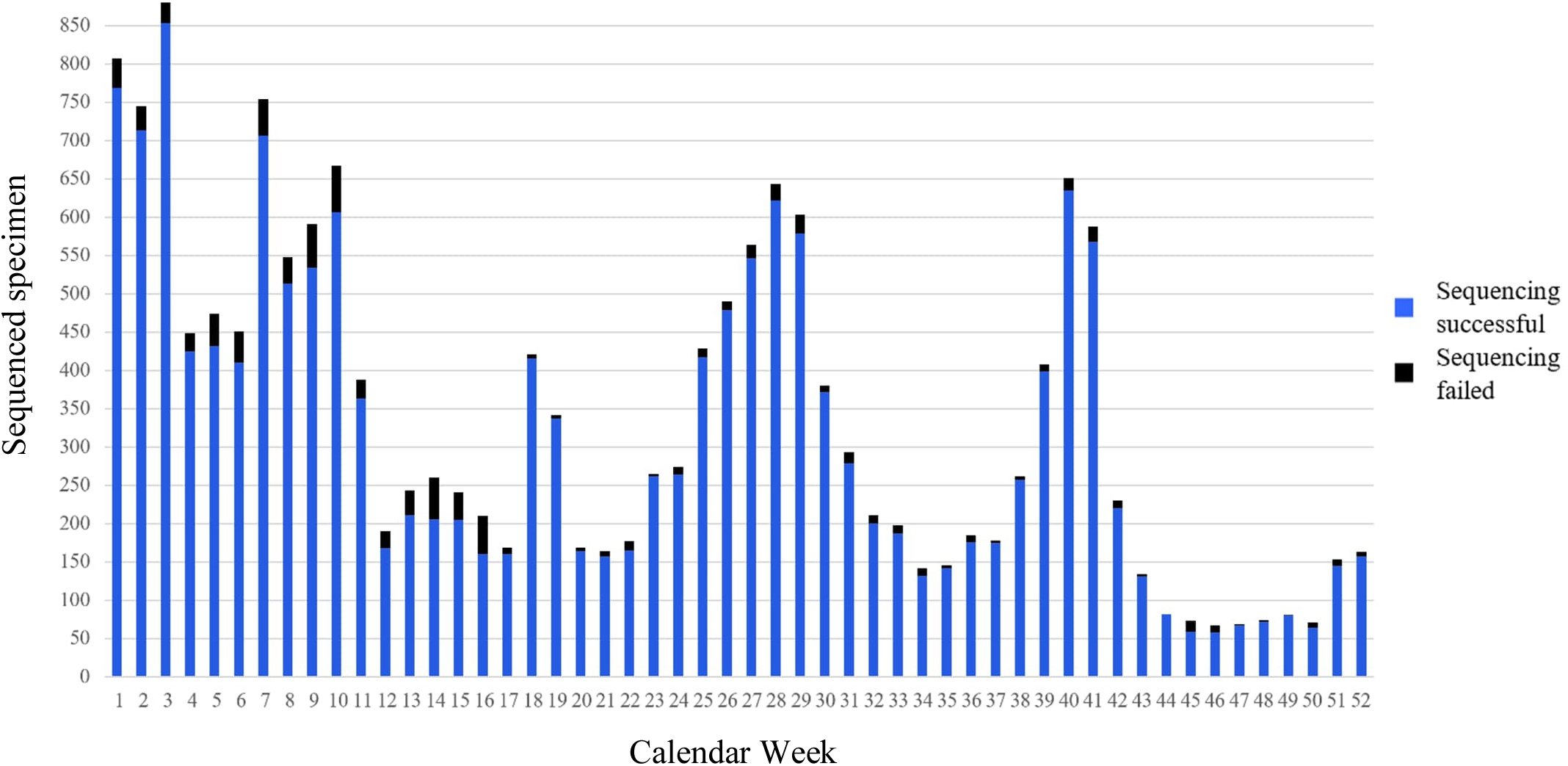
Overview of the number of successful and failed sequencings for Rhine-Neckar region specimen per calendar week for 2022.

A comprehensive overview of the variant distribution in the Rhein-Neckar/Heidelberg region for the year 2022 is presented in Figure 4.

**Figure 4.**
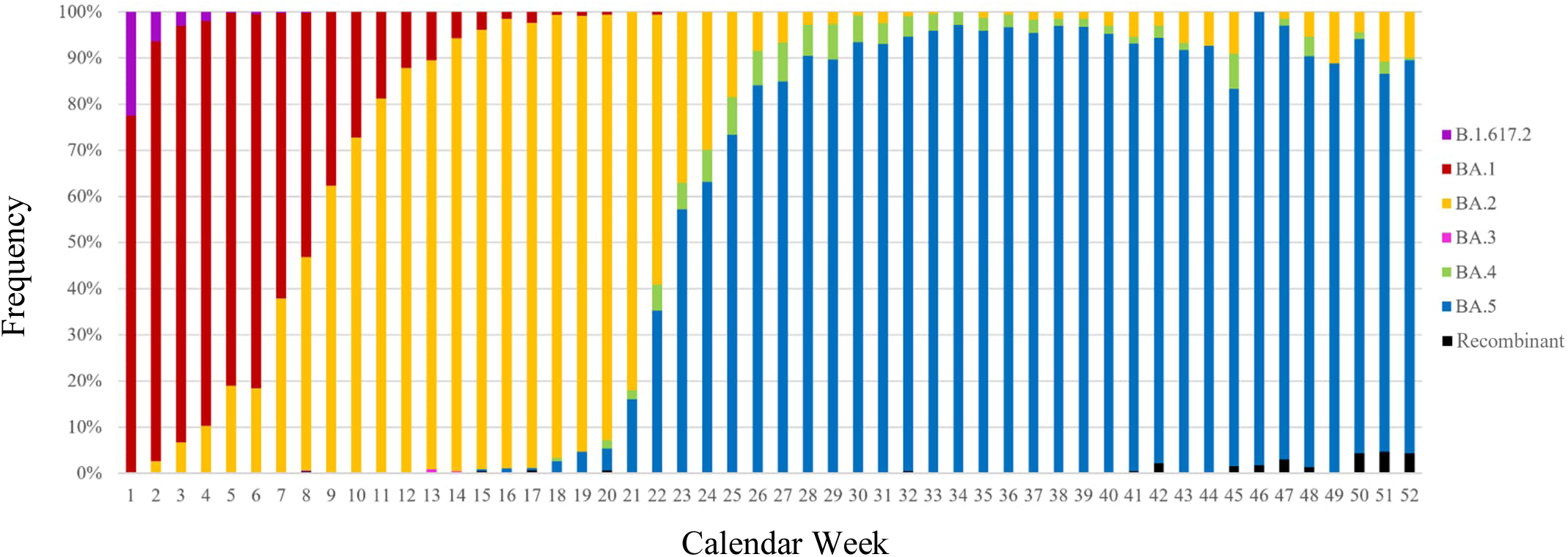
SARS-CoV-2 variant distribution per calendar week 2022.

### Overview of Variant Distribution for the Year 2023 (until July)

The year 2023 started out, like the 2022 ended, with an increasing number of recombinant variants, whereas BA.2(.75) remained widely constant and the cases of BA.5 continued to decrease. These trends continued until calendar week 18, which marked the last week with an identification of a non-recombinant BA.5 and then from calendar week 20 onwards, the recombinant versions where the only variants present (aside from calendar week 26 with 2 specimen with BA.2.75).

With the low SARS-CoV-2 positive PCR specimen numbers since the start of 2023 (although retrospectively the decreasing specimen numbers became already apparent in calendar week 42/2022), which hit rock bottom from calendar week 17 onwards with less than 20 specimen per week until the end of the surveillance period (end of July 2023) our group started to transfer the ARTIC protocol from the NextSeq 500 to the MiSeqDX platform at the end of March 2023.

On May 5, 2023, the WHO declared the end of the SARS-CoV-2 pandemic after more than 3 years and with the end of May 2023 the DESH platform (Robert Koch Institute (RKI), 2021a) was officially discontinued.

An overview of the sequenced specimen for the Rhine-Neckar region in 2023 (until July) is shown in Table 6. The case number reflects the respective variant’s sequence frequencies which fulfilled the RKI DESH criteria (Robert Koch Institute (RKI), 2021b) over the year 2021, the numbers in brackets reflect the total number of specimen which could be assigned to the respective variant following the sequencing process (including samples which passed and failed the RKI DESH criteria). The first and last discovery reflect the date of the respective variant’s first and last detection during the year 2023.

**Table 6.** Variant Distribution in the Rhein-Neckar/Heidelberg Region in 2023.

| Variant | Cases | First Discovery | Last Discovery |
| --- | --- | --- | --- |
| BA.2 | 182 (197) | 01.01.2023 | 30.06.2023 |
| BA.3 | 0 (2) | 18.04.2023 | 23.04.2023 |
| BA.4 | 1 (1) | 04.01.2023 | 04.01.2023 |
| BA.5 | 482 (508) | 01.01.2023 | 04.05.2023 |
| Recombinant | 534 (598) | 02.01.2023 | 31.07.2023 |
| Total | 1199 (1306) |  |  |

In 2023, our group sequenced 1,345 SARS-CoV-2 positive specimen from the Rhine-Neckar/Heidelberg region and as shown in Table 6 for 1,306 samples the lineage and clade were determinable (97.1 percent), leaving only 39 specimens unidentifiable (2.9 percent). 1,199 samples passed the minimal sequencing criteria of the RKI DESH (89.1 percent), which leaves 146 specimen that did not (10.9 percent).

An overview of the number of sequenced samples per calendar week and the success rate of sequencing is depicted in Figure 5.

**Figure 5.**
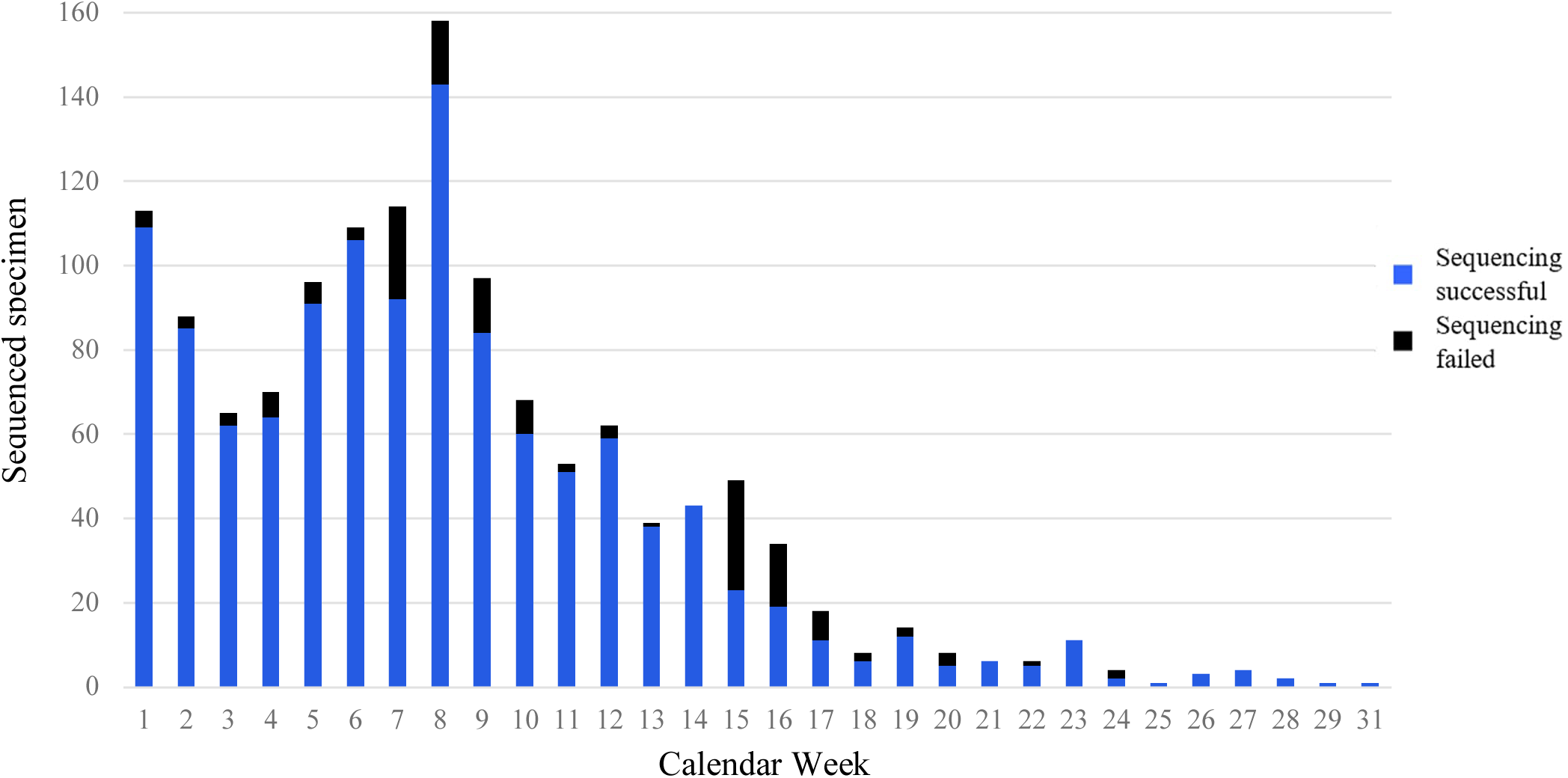
Overview of the number of successful and failed sequencings for Rhine-Neckar region specimen per calendar week for 2023.

A comprehensive overview of the variant distribution in the Rhein-Neckar/Heidelberg region for the year 2023 is presented in Figure 6.

**Figure 6.**
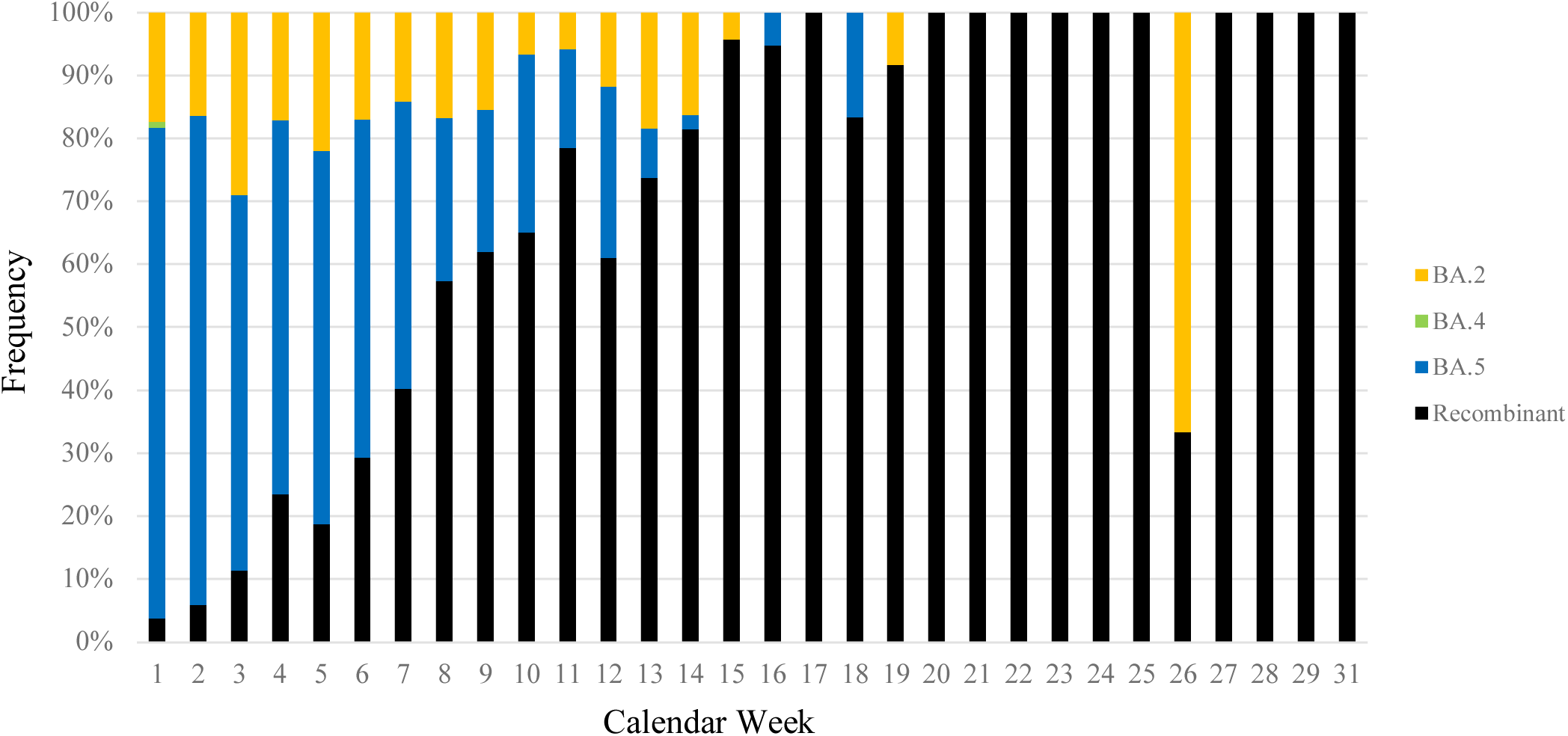
SARS-CoV-2 variant distribution per calendar week 2023.

## Conclusion

In summary, throughout this project we sequenced a wide variety of SARS-CoV-2 variants with significant differences in their epidemiological relevance. While some variants were restricted to specific geographical regions (Mallm et al., 2021), others, such as B.1.617.2 and B.1.1.529, emerged worldwide.

It is likely that the current state of B.1.1.529 with its currently circulating XBB recombinations is not the final state regarding circulating SARS-CoV-2 variants (or mutations) with significant selective advantages over established variants. Therefore, new variants with even greater epidemiological impact will most likely emerge.

The ongoing evolution and emergence of SARS-CoV-2 variants poses ongoing challenges to public health efforts, requiring continuous monitoring, research, and adaptability in response to changing viral dynamics (although the significant reduction of symptom severity and reduction of detection levels over the last few months, put the monitoring requirement into a new perspective). Yet, vigilance and preparedness is still crucial regarding prospective new SARS-CoV-2 variants with a currently undetermined hazard rate regarding their severity and transmissibility to mitigate the potential impact of new variants on public health.

At the end of July 2023 EG.5 started to receive increasing media coverage. Until the end of July 2023 we found not a single EG.5 specimen in the Rhine-Neckar/Heidelberg region (although we detected the first EG.5 case in a specimen from 01/08/2023).

Therefore, the continued and comprehensive surveillance of emerging SARS-CoV-2 strains and mutations by unbiased whole genome sequencing remains of utmost importance for monitoring the still present evolutionary changes in SARS-CoV-2 variants (even after the end of the pandemic).

A complete overview for the evolutionary omicron timeline is shown in Table 7.

**Table 7.** Omicron Distribution in the Rhein-Neckar/Heidelberg Region.

| Variant | Cases | First Discovery | Last Discovery |
| --- | --- | --- | --- |
| BA.1 | 4473 (4714) | 03/12/2021 | 02/09/2022 |
| BA.2 | 4671 (5000) | 07/01/2022 | 30/06/2023 |
| BA.3 | 5 (8) | 22/02/2022 | 23/04/2023 |
| BA.4 | 375 (383) | 02/05/2022 | 04/01/2023 |
| BA.5 | 7546 (7732) | 14/04/2022 | 04/05/2023 |
| Recombinant | 569 (636) | 22/02/2022 | 31/07/2023 |
| Total | 17639 (18473) | 03/12/2021 | 31/07/2023 |

## Limitations

The most significant limitation of this study arises from the fact that our laboratory was not the only sequencing institute in the Rhein-Neckar/Heidelberg region. This aspect was already addressed in 2021 by comparing the variant distribution (or sequencing results) of this study for each calendar week with the sequencing results from the Limbach Laboratory Heidelberg, which showed no significant variation of the circulating variants (Differences of more than 10 percent in individual variant frequencies for at least two consecutive weeks would have been considered as statistically significant).

## Acknowledgments

We thank Christian Drosten and Barbara Mühlemann from the German National Consultant Laboratory for Coronaviruses for discussion on SARS-CoV-2 variant distribution. We gratefully acknowledge the help of Maria Anders-Össwein with serological analyses and of Marie Bartenschlager with the neutralization assays, and thank all diagnostics employees and EMBL GeneCore’s NGS protocol-automation team for their support. This work was supported by the program for surveillance and control of SARS-CoV-2 mutations of the state of Baden-Württemberg, the German Federal Research Network Applied Surveillance and Testing (B-FAST) within the Network University Medicine and the DKFZ@fightCOVID initiative.

## Author’s contributions

Conceptualization: HGK, KR, RB

Investigation: CB, JPM, HK, NW, JK, KBö, KBa, KLJ, SP, BM, VB, NA, AT

Data Curation: CB, NW, DH, TR

Formal Analysis: TR, VB, SS, IL, TR, JPM, HK, KR

Writing – Original Draft Preparation: CB

Writing – Review & Editing: PS, NW, HGK, KR, JPM, RB, BM, KB, CB, TR, VB, MB

Supervision: PS, HGK, KR, RB, MB

## Appendix A

**Table A1.** Non-VOC statistics for the Rhine-Neckar region.

| Variant | Cases | First Discovery | Last Discovery | Clade |
| --- | --- | --- | --- | --- |
| A.5 | 0 (1) | 29/01/2021 | 29/01/2021 | 19B |
| A.29 | 2 (7) | 31/01/2021 | 24/04/2021 | 19B |
| AA.1 | 2 (3) | 14/01/2021 | 10/03/2021 | 20E |
| B.1 | 9 (20) | 17/01/2021 | 29/05/2021 | 20A/20B/20E |
| B.1.1 | 24 (32) | 12/01/2021 | 02/05/2021 | 20B |
| B.1.1.28 | 2 (2) | 25/01/2021 | 25/01/2021 | 20B |
| B.1.1.39 | 22 (23) | 13/01/2021 | 24/02/2021 | 20B |
| B.1.1.70 | 13 (14) | 12/01/2021 | 14/02/2021 | 20B |
| B.1.1.153 | 3 (4) | 19/01/2021 | 30/01/2021 | 20B |
| B.1.1.161 | 0 (1) | 13/01/2021 | 13/01/2021 | 20B |
| B.1.1.170 | 27 (28) | 08/01/2021 | 20/03/2021 | 20B |
| B.1.1.189 | 6 (6) | 05/01/2021 | 22/01/2021 | 20B |
| B.1.1.232 | 2 (2) | 12/01/2021 | 20/01/2021 | 20B |
| B.1.1.236 | 13 (14) | 13/01/2021 | 27/01/2021 | 20B |
| B.1.1.277 | 1 (1) | 15/01/2021 | 15/01/2021 | 20B |
| B.1.1.294 | 1 (1) | 29/01/2021 | 29/01/2021 | 20B |
| B.1.1.297 | 24 (28) | 04/01/2021 | 01/03/2021 | 20B |
| B.1.1.317 | 7 (7) | 13/01/2021 | 12/03/2021 | 20B |
| B.1.1.318 | 2 (2) | 03/05/2021 | 03/05/2021 | 20B |
| B.1.1.347 | 1 (1) | 19/01/2021 | 19/01/2021 | 20B |
| B.1.1.374 | 2 (2) | 04/02/2021 | 28/02/2021 | 20B |
| B.1.1.385 | 38 (38) | 14/01/2021 | 25/02/2021 | 20B |
| B.1.1.519 | 9 (9) | 01/03/2021 | 28/03/2021 | 20B |
| B.1.2 | 8 (9) | 19/01/2021 | 15/02/2021 | 20G |
| B.1.9 | 12 (14) | 03/01/2021 | 26/03/2021 | 20A |
| B.1.36 | 4 (4) | 21/01/2021 | 15/02/2021 | 20A |
| B.1.160 | 106 (111) | 02/01/2021 | 22/10/2021 | 20A |
| B.1.177 | 143 (154) | 06/01/2021 | 25/03/2021 | 20E |
| B.1.221 | 76 (83) | 05/01/2021 | 13/04/2021 | 20A |
| B.1.222 | 0 (1) | 14/01/2021 | 14/01/2021 | 20A |
| B.1.258 | 129 (135) | 04/01/2021 | 23/03/2021 | 20A |
| B.1.406 | 4 (4) | 23/01/2021 | 05/02/2021 | 20A |
| B.1.474 | 1 (1) | 23/01/2021 | 23/01/2021 | 20A |
| B.1.579 | 0 (1) | 20/01/2021 | 20/01/2021 | 20A |
| B.1.619 | 6 (7) | 03/05/2021 | 25/06/2021 | 20A |
| B.1.621 | 1 (1) | 14/06/2021 | 14/06/2021 | 21H |
| B.55 | 0 (1) | 22/01/2021 | 22/01/2021 | 19A |
| C.35 | 9 (9) | 13/01/2021 | 08/02/2021 | 20D |
| C.36 | 8 (8) | 14/ 01/2021 | 26/06/2021 | 20D |
| C.38 | 1 (1) | 16/05/2021 | 16/05/2021 | 20D |
| G.1 | 1 (1) | 21/01/2021 | 21/01/2021 | 20A |
| R.1 | 3 (3) | 26/02/2021 | 28/02/2021 | 20B |
| Total | 722 (794) | 02/01/2021 | 26/06/2021 |  |

**Table A2.**
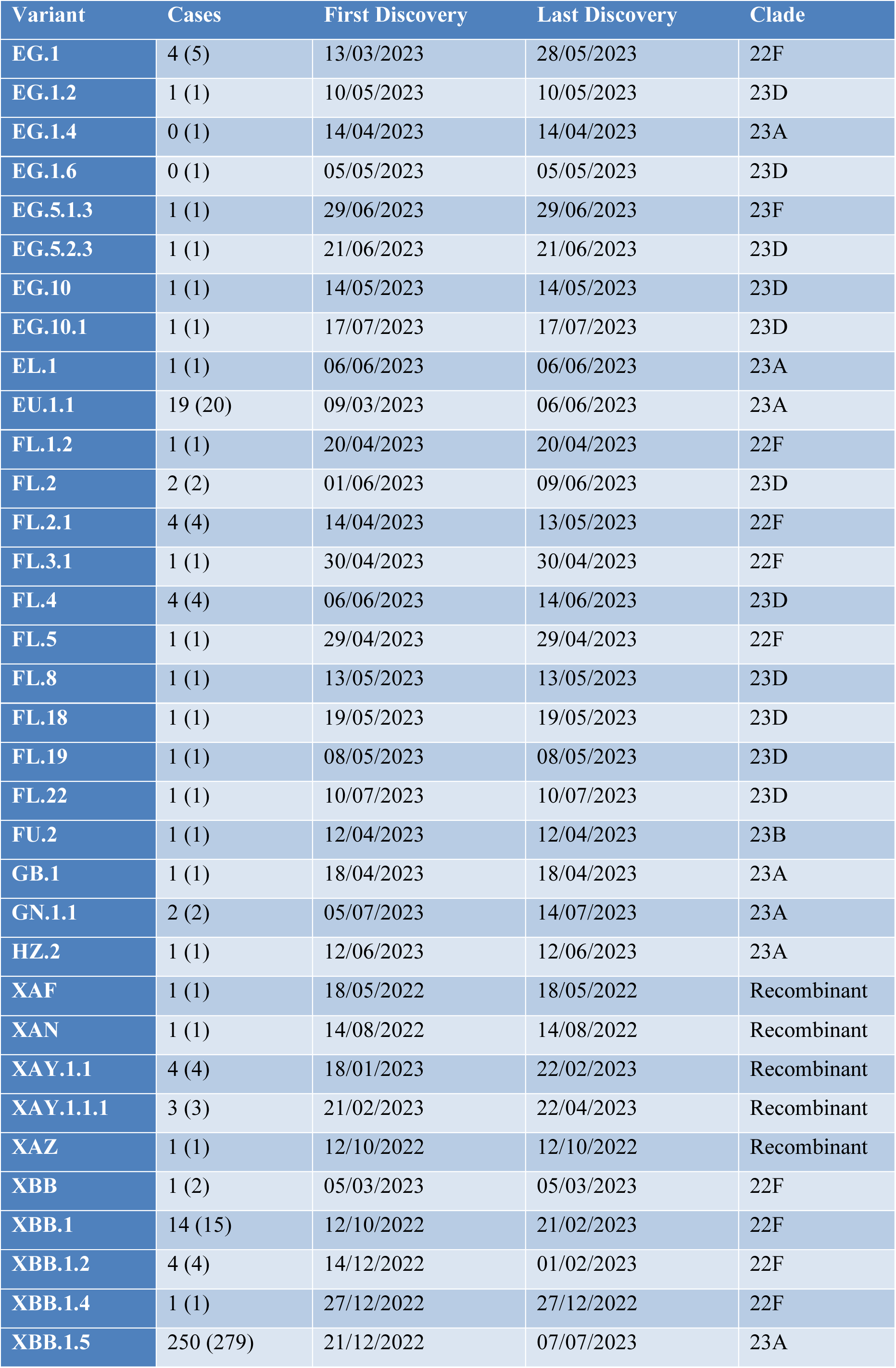
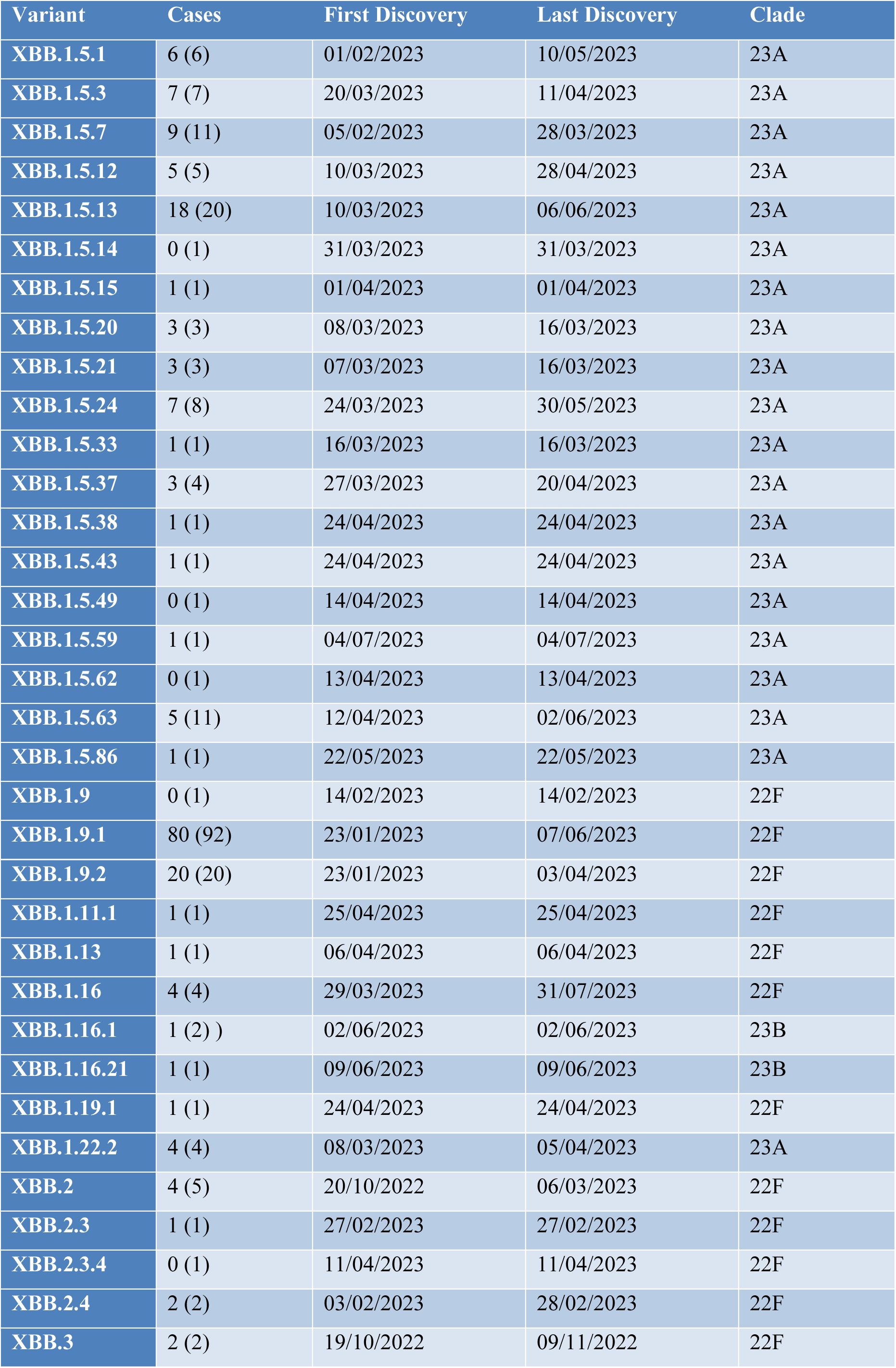
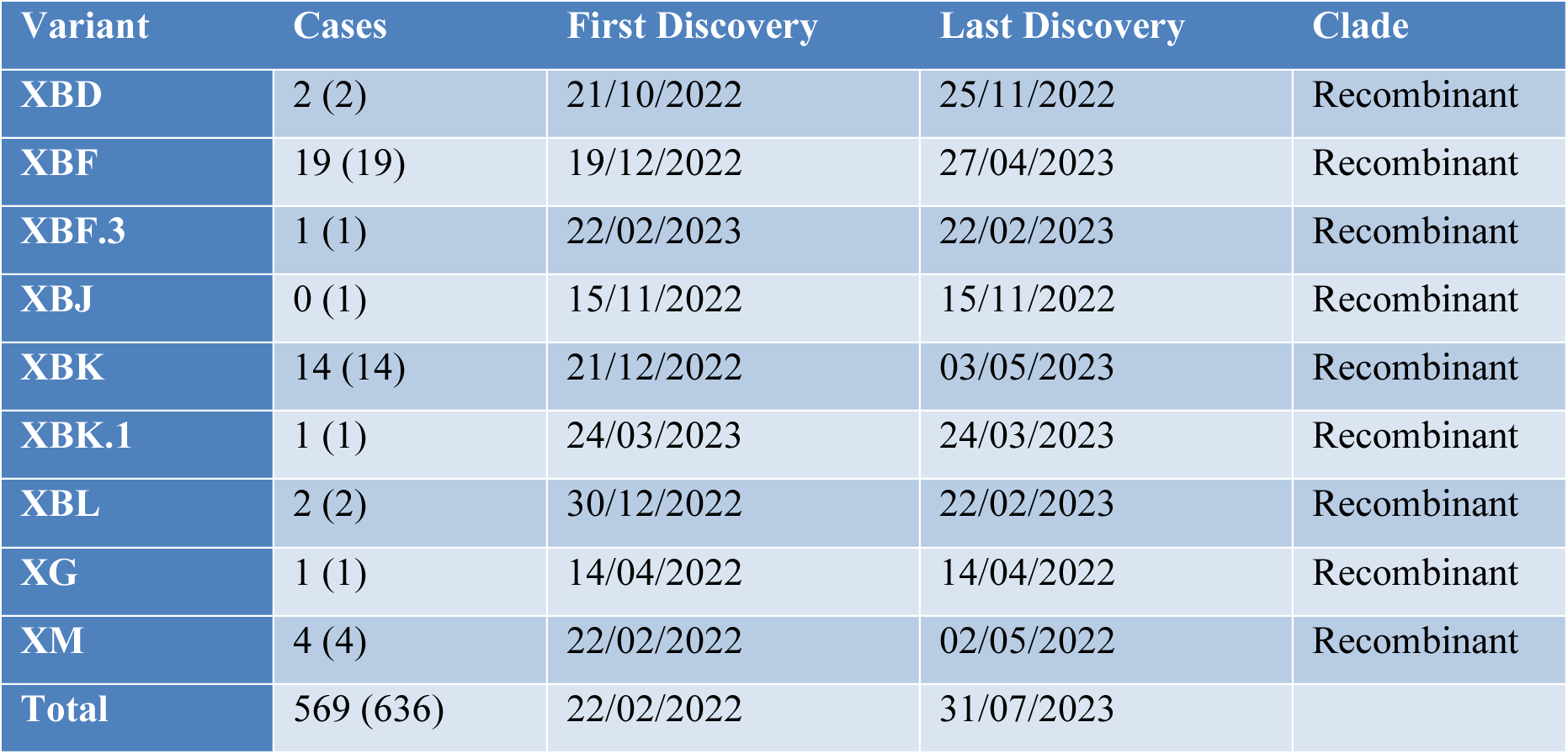
Recombinant variant(s) statistics for the Rhine-Neckar region.

## Appendix B ARTIC SARS-CoV-2 protocol (Tyson et al., 2020), currently V4

The NEBNext ARTIC SARS-CoV-2 FS Library Prep Kit for Illumina (E7650) contains the enzymes, buffers and oligonucleotides required to convert a broad range of total RNA into high quality, targeted, libraries for next-generation sequencing on the Illumina platform. Primers targeting the human EDF1 (NEBNext ARTIC Human Primer Mix 1) and NEDD8 (NEBNext ARTIC Human Primer Mix 2) genes are supplied as optional internal controls. The fast, user-friendly workflow also has minimal hands-on time. Each kit component must pass rigorous quality control standards, and for each new lot the entire set of reagents is functionally validated together by construction and sequencing of indexed libraries on an Illumina sequencing platform. For larger volume requirements, customized and bulk packaging is available by purchasing through the OEM/Bulks department at NEB.

**Figure 1.**
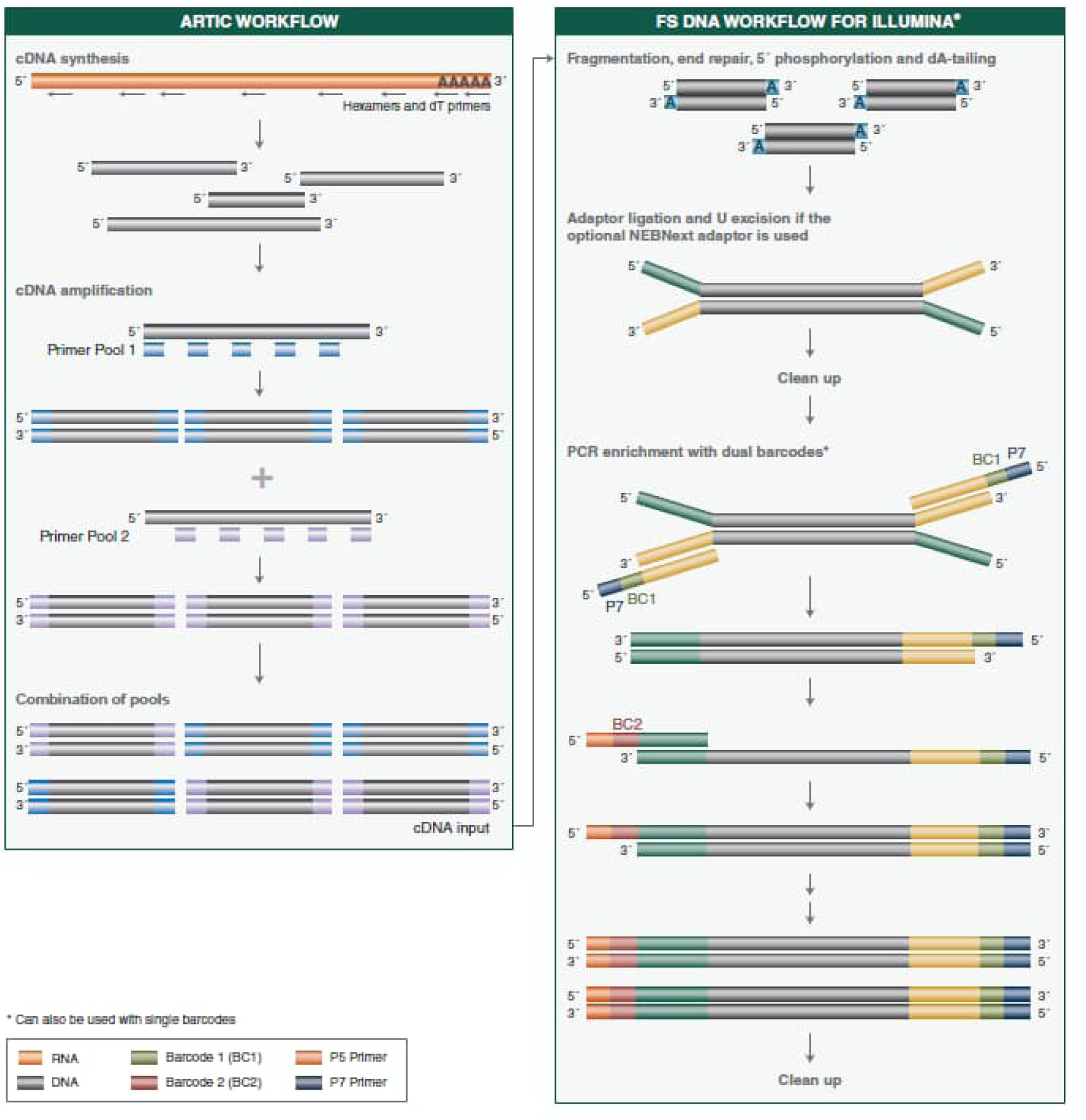
Workflow demonstrating the use of NEBNext ARTIC SARS-coV-2 FS Library Prep Kit.

### 1. cDNA Synthesis

The presence of carry-over products can interfere with sequencing accuracy, particularly for low copy targets. Therefore, it is important to carry out the appropriate no template control (NTC) reactions to demonstrate that positive reactions are meaningful.

1.1. Gently mix and spin down the LunaScript RT SuperMix reagent. Prepare the cDNA synthesis reaction as described below:

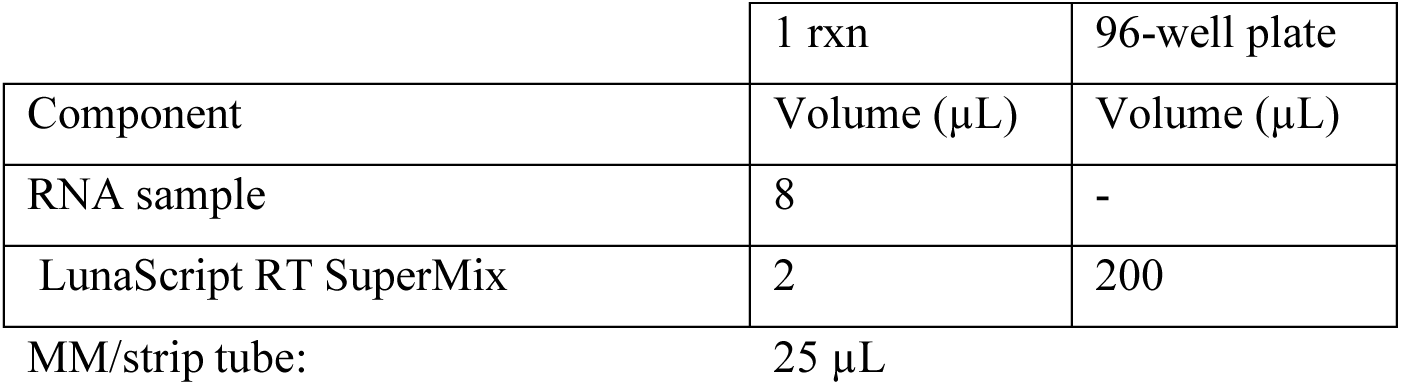

|  | 1 rxn | 96-well plate |
| --- | --- | --- |
| Component | Volume (μL) | Volume (μL) |
| RNA sample | 8 | - |
| LunaScript RT SuperMix | 2 | 200 |

MM/strip tube: 25 μL

❖ For no template controls, add water instead of RNA.
❖ Each RNA plate should have a no template (H_2_O), a Human RNA sample and Twist RNA (12500 copies) as controls.
❖ The wells to add the controls should be constant:

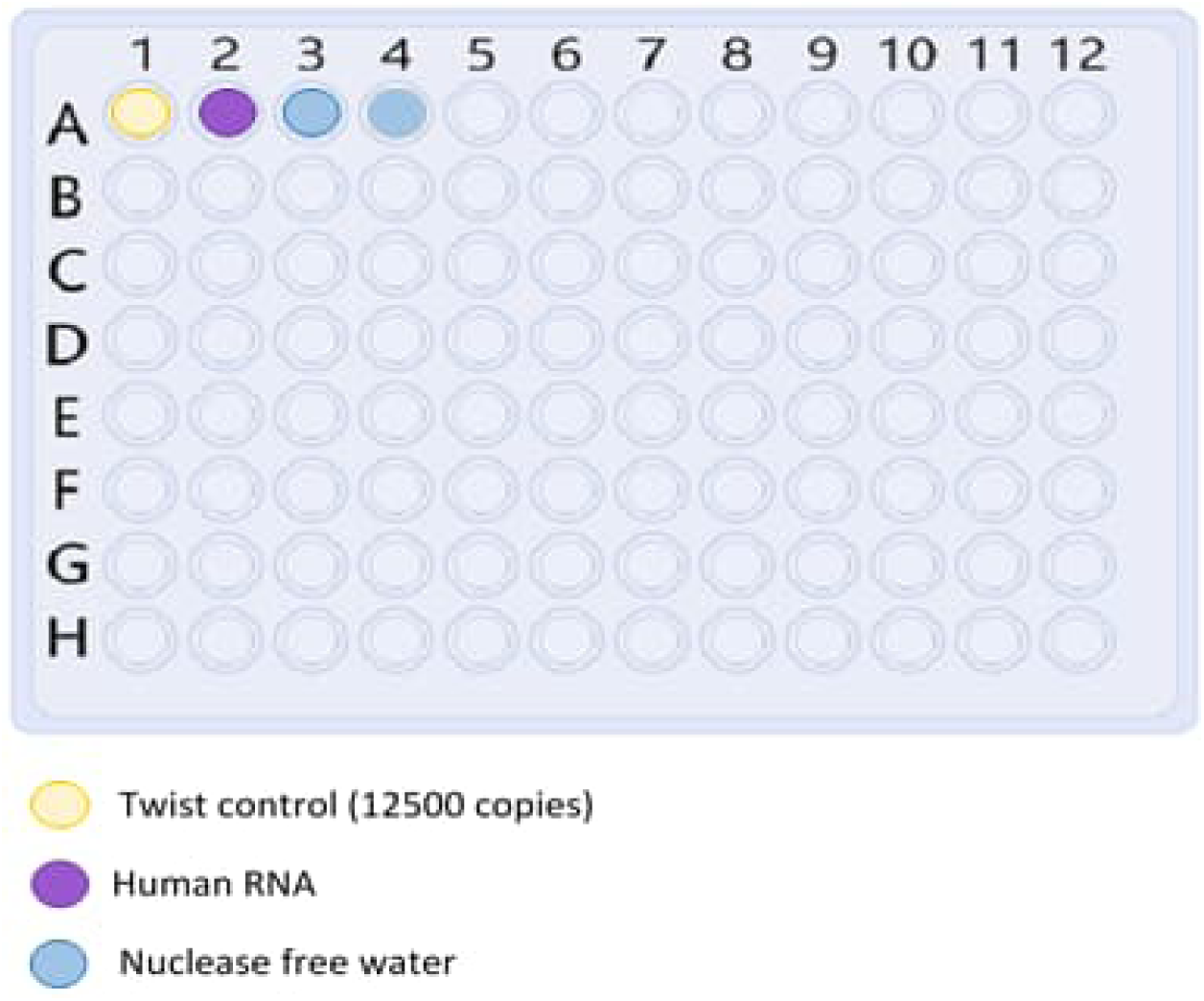

The controls should be switched each plate!

1.2. Seal the plate, vortex and spin prior to placing it in the thermocycler.

1.3. Incubate the reactions in a thermocycler with the following steps:

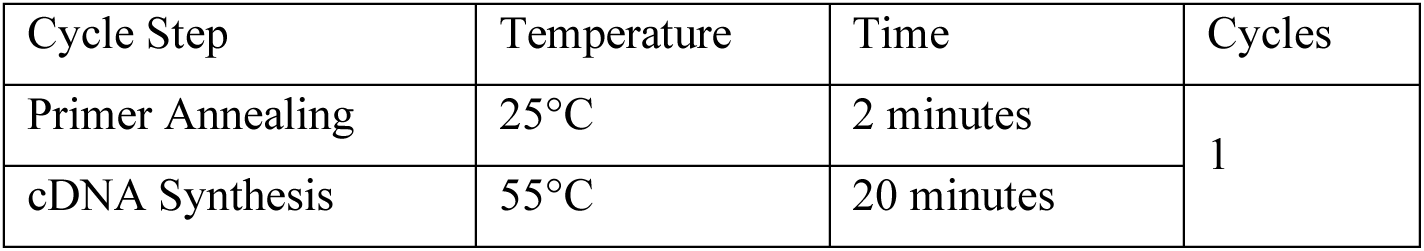
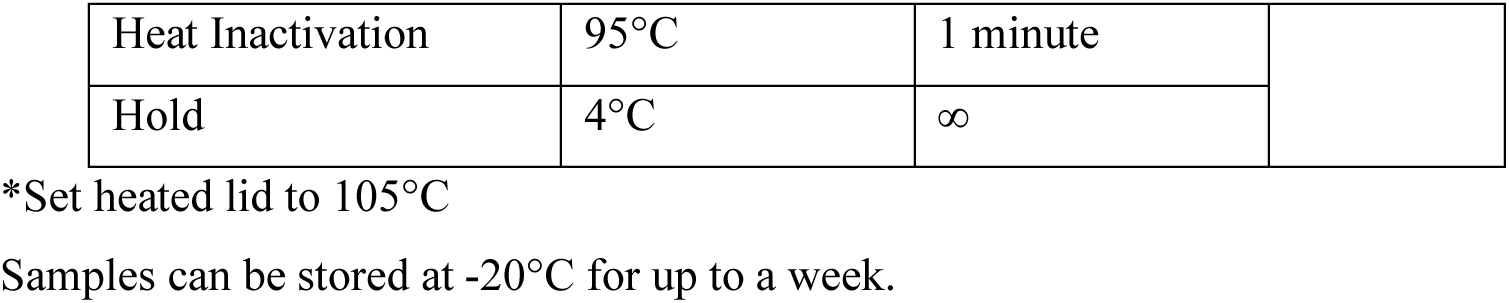

| Cycle Step | Temperature | Time | Cycles |
| --- | --- | --- | --- |
| Primer Annealing | 25°C | 2 minutes | 1 |
| cDNA Synthesis | 55°C | 20 minutes |  |
| Heat Inactivation | 95°C | 1 minute |  |
| Hold | 4°C | ∞ |  |

\*Set heated lid to 105°C
Samples can be stored at -20°C for up to a week.

### 2. cDNA Amplification

**Note:** 4.5 μl cDNA input is recommended. If using less than 4.5 μl of cDNA, add nuclease-free water to a final volume of 4.5 μl. We recommend setting up the cDNA synthesis and cDNA amplification reactions in different rooms to minimize cross-contamination of future reactions.

2.1. Gently mix and spin down reagents. Prepare the split pool cDNA amplification reactions as described below:

For Pool Set A:

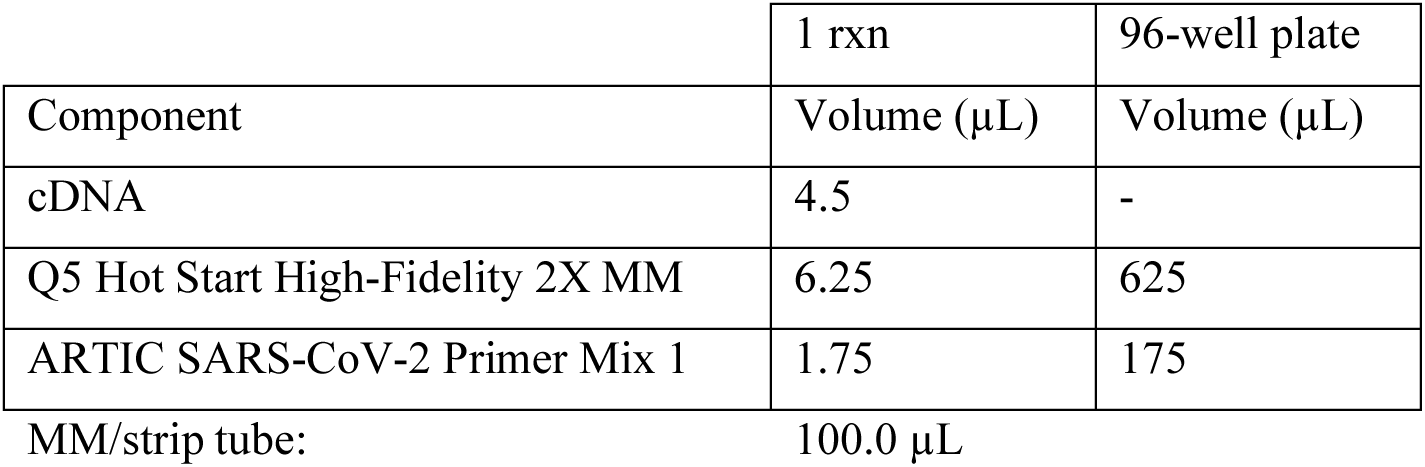

For Pool Set B:

MM/strip tube: 100.0 µL For Pool Set B:
|  | 1 rxn | 96-well plate |
| --- | --- | --- |
| Component | Volume (µL) | Volume (µL) |
| cDNA | 4.5 | - |
| Q5 Hot Start High-Fidelity 2X MM | 6.25 | 625 |
| ARTIC SARS-CoV-2 Primer Mix 2 | 1.75 | 175 |

MM/strip tube: 100.0 µL

2.2. Seal the plate, vortex and spin prior to placing it in the thermocycler.

2.3. Incubate reactions in a thermocycler with the following steps:

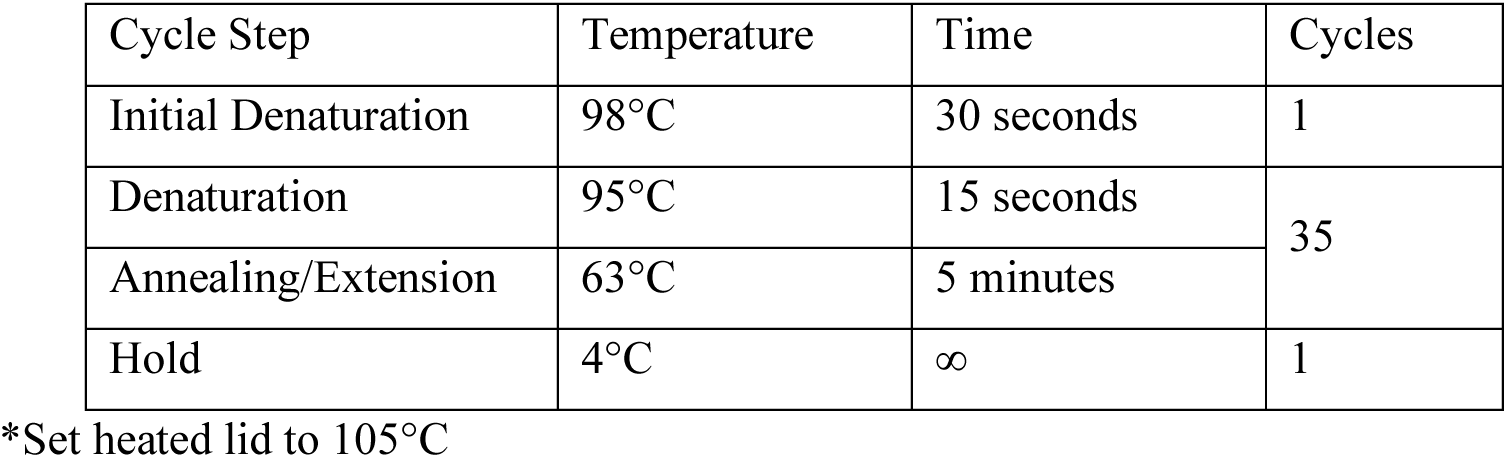

2.4. Combine the Pool A and Pool B PCR reactions for each sample.

2.5. Quality control

We used to perform Qubit as well as bioanalyzer, but we found that Qubit alone was predictive of whether libraries would succeed. We measured the controls and 8 random samples (2 samples from each source plate) with Qubit.

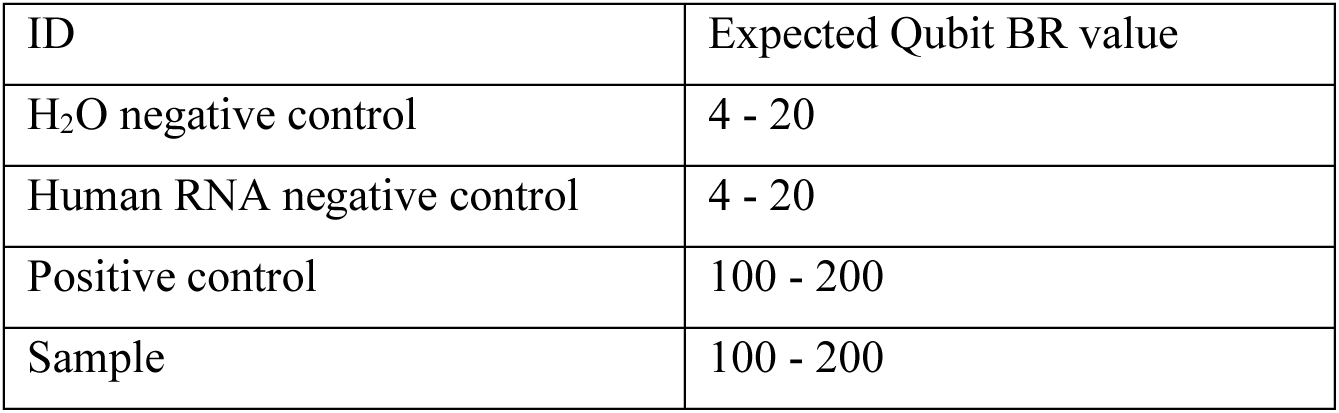

Samples can be stored at −20°C for up to a week.

### 3. Fragmentation/End Prep

3.1. Mix 1.3 μl of the pooled amplicons (from step 2.4) with 11.7 μl of nuclease free water in a new 96well plate (Eppendorf TwinPlate) – Store the rest of the pools at –20°C

3.2. Ensure that the Ultra II FS Reaction Buffer is completely thawed. If a precipitate is seen in the buffer, pipette up and down several times to break it up, and quickly vortex to mix. Place on ice until use.

3.3. Vortex the Ultra II FS Enzyme Mix 5-8 seconds prior to use and place on ice.

**Note:** It is important to vortex the enzyme mix prior to use for optimal performance.

3.4. Add the following components to a 0.2 ml thin wall PCR tube on ice:

|  | 1 rxn | 96-well plate |
| --- | --- | --- |
| Component | Volume (µL) | Volume (µL) |
| cDNA amplified | 13 | - |
| NEBNext Ultra II FS reaction Buffer | 3.5 | 350 |
| NEBNext Ultra II FS Enzyme Mix | 1 | 100 |

MM/strip tube: 56.2 µL

3.5. Seal the plate, vortex and spin prior to placing it in the thermocycler.

3.6. In a thermocycler, run the following program:

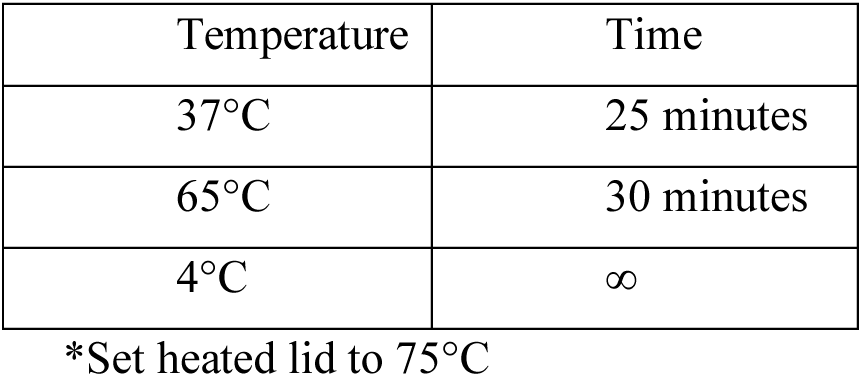

If necessary, samples can be stored at –20°C; however, a slight loss in yield (∼20%) may be observed. We recommend continuing with adaptor ligation before stopping.

### 4. Adaptor Ligation

4.1. Add the following components directly to the FS Reaction Mixture:

|  | 1 rxn | 96-well plate |
| --- | --- | --- |
| Component | Volume (μL) | Volume (μL) |
| FS reaction Mixture | 17.5 | - |
| NEBNext Adaptor for Illumina | 1.25 | 125 |
| NEBNext Ultra II Ligation MM | 15 | 1500 |

MM/strip tube: 203.1 μL
\*Mix the Ultra II Ligation Master Mix by pipetting up and down several times prior to adding to the reaction.
\*\*The NEBNext adaptor is provided in NEBNext Oligo kits.

4.2. Seal the plate, vortex and spin prior to placing it in the thermocycler

4.3. Incubate at 20°C for 15 minutes in a thermocycler with the heated lid off.

4.4. Add 1.5 μl of • (red) USER® Enzyme to the ligation mixture from Step 4.3.

4.5. Mix well and incubate at 37°C for 15 minutes with the heated lid set to ≥ 47°C. Samples can be stored overnight at –20°C.

### 5. PCR Enrichment of Adaptor-ligated DNA/Dual Index PCR

5.1. Add the following components to a sterile strip tube:

|  | 1 rxn | 96-well plate |
| --- | --- | --- |
| Component | Volume (μL) | Volume (μL) |
| Adaptor Ligated DNA Fragments | 7.5 | - |
| NEBNext Library PCR MM | 12.5 | 1250 |
| Dual Index Mix | 5 | - |

MM/strip tube: 156.2 μl
\*Oligos from NEB dual index 96-well plate. 4 different sets.
\*\*The leftover of the Adaptor Ligated DNA Fragments can be stored overnight @ –20°C.

5.2. Seal the plate, vortex and spin before starting the program in the thermocycler.

5.3. Place the tube on a thermocycler and perform PCR amplification using the following PCR cycling Conditions:

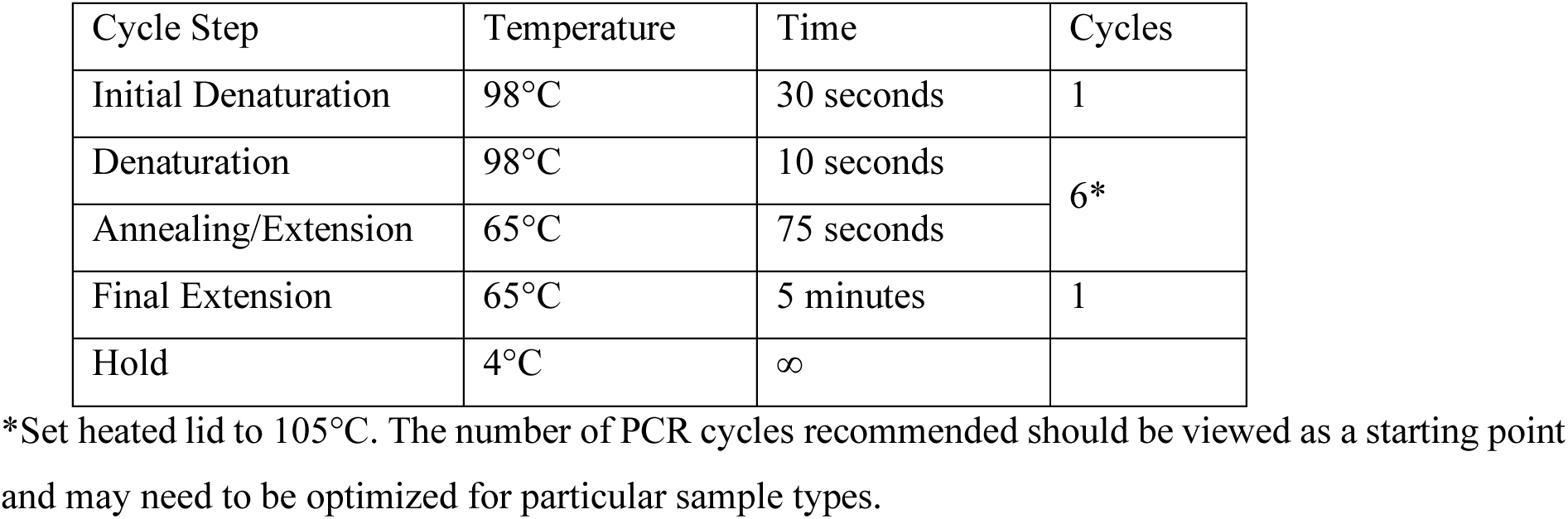

### 6. Pooling of libraries

6.1. Without any QC step, all libraries from a plate should be pooled together.

6.2. Pipette 4 µL of each library into a strip with a multichannel pipette and combine all volume into a 1.5mL Eppendorf tube (∼380 µL).

### 7. Cleanup of the pool

7.1. Vortex NEBNext Sample Purification Beads to resuspend.

7.2. Measure the pool volume and add 0.7X of resuspended beads to the PCR reaction. Mix well by pipetting up and down at least 10 times. Be careful to expel all of the liquid out of the tip during the last mix. Vortexing for 3-5 seconds on high can also be used. If centrifuging samples after mixing, be sure to stop the centrifugation before the beads start to settle out.

7.3. Incubate samples on bench top for at least 5 minutes at room temperature.

7.4. Place the tube/plate on an appropriate magnetic stand to separate the beads from the supernatant. If necessary, quickly spin the sample to collect the liquid from the sides of the tube or plate wells before placing on the magnetic stand.

7.5. After 5 minutes (or when the solution is clear), carefully remove and discard the supernatant. Be careful not to disturb the beads that contain DNA targets.

**Note:** do not discard the beads.

7.6. Add 200 μl of 80% freshly prepared ethanol to the tube/plate while in the magnetic stand. Incubate at room temperature for 30 seconds, and then carefully remove and discard the supernatant. Be careful not to disturb the beads that contain DNA targets.

7.7. Repeat Step 7.6. for a total of two washes. Be sure to remove all visible liquid after the second wash. If necessary, briefly spin the tube/plate, place back on the magnet and remove traces of ethanol with a p10 pipette tip.

7.8. Air dry the beads for up to 5 minutes while the tube/plate is on the magnetic stand with the lid open.

**Note:** Do not over-dry the beads. This may result in lower recovery of DNA.

7.8.1. Remove the tube/plate from the magnetic stand. Elute the DNA target from the beads by adding 105 μl of 0.1x TE. Mix well by pipetting up and down 10 times, or on a vortex mixer. Incubate for at least 2 minutes at room temperature.

7.8.2. If necessary, quickly spin the sample to collect the liquid from the sides of the tube or plate wells before placing back on the magnetic stand.

7.8.3. Place the tube/plate on the magnetic stand. After 5 minutes (or when the solution is clear), transfer 100 μl to a new eppendorf tube.

7.9. Repeat the cleanup steps with the 0.9x of resuspended beads

7.10. Elute the DNA target from the beads by adding 55 μl of 0.1x TE and mix well

7.11. Place the tube/plate on the magnetic stand. After 5 minutes (or when the solution is clear), transfer 50 μl to a new eppendorf tube and store at –20°C.

### 8. Library QC

8.1. Measure the concentration of the final pool with Qubit DNA BR kit.

8.2. Assess the library size distribution with Agilent Bioanalyzer high sensitivity DNA reagents. The sample may need to be highly diluted.

8.3. The expected concentration is between 100 and 200 ng/ul by BR Qubit.

8.4. A peek sized of 200–250 bp is expected on a Bioanalyzer, based on a 30-minute fragmentation time:

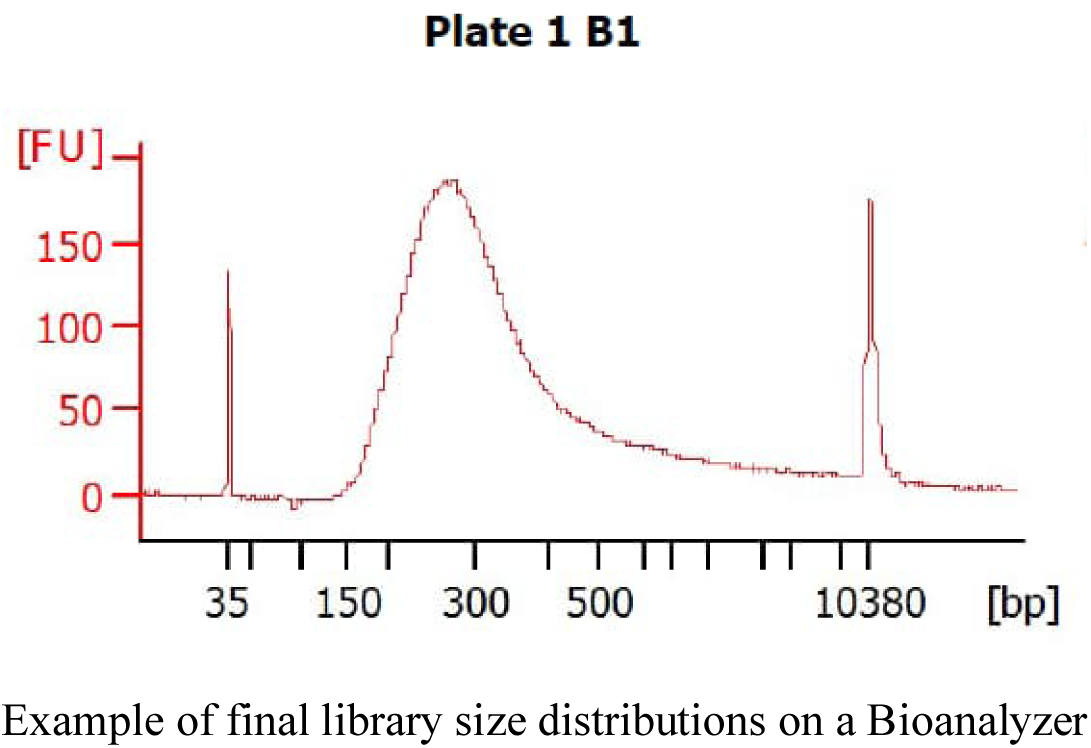

## References

1. Abdool Karim, S. S. & de Oliveira, T. 2021. New SARS-CoV-2 Variants - Clinical, Public Health, and Vaccine Implications. N Engl J Med, published online 24 March 2021.

2. Center for Systems Science and Engineering (Johns Hopkins University). 2023. COVID-19 Dashboard [Online]. Available: https://coronavirus.jhu.edu/map.html [Accessed 19/01/2023].

3. Clarke, K. E. N., Jones, J. M., Deng, Y., Nycz, E., Lee, A., Iachan, R., Gundlapalli, A. V., Hall, A. J. & MacNeil, A. 2022. Seroprevalence of Infection-Induced SARS-CoV-2 Antibodies - United States, September 2021-February 2022. MMWR Morb Mortal Wkly Rep, 71, 606–608.

4. Ewels, P. A., Peltzer, A., Fillinger, S., Patel, H., Alneberg, J., Wilm, A., Garcia, M. U., Di Tommaso, P. & Nahnsen, S. 2020. The nf-core framework for community-curated bioinformatics pipelines. Nat Biotechnol, 38, 276–278.

5. Garrison, E. & Marth, G. 2012. Haplotype-based variant detection from short-read sequencing. arXiv, 1207.

6. Grubaugh, N. D., Gangavarapu, K., Quick, J., Matteson, N. L., De Jesus, J. G., Main, B. J., Tan, A. L., Paul, L. M., Brackney, D. E., Grewal, S., Gurfield, N., Van Rompay, K. K. A., Isern, S., Michael, S. F., Coffey, L. L., Loman, N. J. & Andersen, K. G. 2019. An amplicon-based sequencing framework for accurately measuring intrahost virus diversity using PrimalSeq and iVar. Genome Biol, 20, 8.

7. Hadfield, J., Megill, C., Bell, S. M., Huddleston, J., Potter, B., Callender, C., Sagulenko, P., Bedford, T. & Neher, R. A. 2018. Nextstrain: real-time tracking of pathogen evolution. Bioinformatics, 34, 4121–4123.

8. Havers, F. P., Reed, C., Lim, T., Montgomery, J. M., Klena, J. D., Hall, A. J., Fry, A. M., Cannon, D. L., Chiang, C. F., Gibbons, A., Krapiunaya, I., Morales-Betoulle, M., Roguski, K., Rasheed, M. A. U., Freeman, B., Lester, S., Mills, L., Carroll, D. S., Owen, S. M., Johnson, J. A., Semenova, V., Blackmore, C., Blog, D., Chai, S. J., Dunn, A., Hand, J., Jain, S., Lindquist, S., Lynfield, R., Pritchard, S., Sokol, T., Sosa, L., Turabelidze, G., Watkins, S. M., Wiesman, J., Williams, R. W., Yendell, S., Schiffer, J. & Thornburg, N. J. 2020. Seroprevalence of Antibodies to SARS-CoV-2 in 10 Sites in the United States, March 23-May 12, 2020. JAMA Intern Med.

9. Li, H. 2011. A statistical framework for SNP calling, mutation discovery, association mapping and population genetical parameter estimation from sequencing data. Bioinformatics, 27, 2987–93.

10. Li, H. & Durbin, R. 2009. Fast and accurate short read alignment with Burrows-Wheeler transform. Bioinformatics, 25, 1754–60.

11. Li, H., Handsaker, B., Wysoker, A., Fennell, T., Ruan, J., Homer, N., Marth, G., Abecasis, G. & Durbin, R. 2009. The Sequence Alignment/Map format and SAMtools. Bioinformatics, 25, 2078–9.

12. Mallm, J.-P., Bundschuh, C., Kim, H., Weidner, N., Steiger, S., Lander, I., Börner, K., Bauer, K., Hübschmann, D., Benes, V., Rausch, T., de Azevedo, N. T. D., Telzerow, A., Jost, K. L., Parthé, S., Schnitzler, P., Boutros, M., Müller, B., Bartenschlager, R., Kräusslich, H.-G. & Rippe, K. 2021. Local emergence and decline of a SARS-CoV-2 variant with mutations L452R and N501Y in the spike protein. medRxiv, 2021.04.27.21254849.

13. McLaren, W., Gil, L., Hunt, S. E., Riat, H. S., Ritchie, G. R., Thormann, A., Flicek, P. & Cunningham, F. 2016. The Ensembl Variant Effect Predictor. Genome Biol, 17, 122.

14. Patel, H. V., S.; Monzón, S.; Espinosa-Carrasco, J.; Heuer, M. L.; Gabernet, G.; Julia, M.; Kelly, S.; Sameith, K.; Garcia, M. 2020. nf-core/viralrecon: nf-core/viralrecon v1.1.0 - Steel Pangolin.

15. Rambaut, A., Holmes, E. C., O’Toole, Á., Hill, V., McCrone, J. T., Ruis, C., du Plessis, L. & Pybus, O. G. 2020. A dynamic nomenclature proposal for SARS-CoV-2 lineages to assist genomic epidemiology. Nat Microbiol, 5, 1403–1407.

16. Rausch, T., Fritz, M. H., Untergasser, A. & Benes, V. 2020. Tracy: basecalling, alignment, assembly and deconvolution of sanger chromatogram trace files. BMC Genomics, 21, 230.

17. Rausch, T., Hsi-Yang Fritz, M., Korbel, J. O. & Benes, V. 2019. Alfred: interactive multi-sample BAM alignment statistics, feature counting and feature annotation for long- and short-read sequencing. Bioinformatics, 35, 2489–2491.

18. Robert Koch Institute (RKI). 2021a. Deutscher Elektronischer Sequenzdaten-Hub (DESH) [Online]. Available: https://www.rki.de/DE/Content/InfAZ/N/Neuartiges_Coronavirus/DESH/DESH.html [Accessed 06/12/2021].

19. Robert Koch Institute (RKI). 2021b. Qualitätsvorgaben für die Sequenzdaten [Online]. Available: https://www.rki.de/DE/Content/InfAZ/N/Neuartiges_Coronavirus/DESH/Qualitaetskriterien.pdf [Accessed 06/12/2021].

20. Shu, Y. & McCauley, J. 2017. GISAID: Global initiative on sharing all influenza data - from vision to reality. Euro Surveill, 22.

21. Stringhini, S., Wisniak, A., Piumatti, G., Azman, A. S., Lauer, S. A., Baysson, H., De Ridder, D., Petrovic, D., Schrempft, S., Marcus, K., Yerly, S., Arm Vernez, I., Keiser, O., Hurst, S., Posfay-Barbe, K. M., Trono, D., Pittet, D., Gétaz, L., Chappuis, F., Eckerle, I., Vuilleumier, N., Meyer, B., Flahault, A., Kaiser, L. & Guessous, I. 2020. Seroprevalence of anti-SARS-CoV-2 IgG antibodies in Geneva, Switzerland (SEROCoV-POP): a population-based study. Lancet, 396, 313–319.

22. Tyson, J. R., James, P., Stoddart, D., Sparks, N., Wickenhagen, A., Hall, G., Choi, J. H., Lapointe, H., Kamelian, K., Smith, A. D., Prystajecky, N., Goodfellow, I., Wilson, S. J., Harrigan, R., Snutch, T. P., Loman, N. J. & Quick, J. 2020. Improvements to the ARTIC multiplex PCR method for SARS-CoV-2 genome sequencing using nanopore. bioRxiv.

23. Wood, D. E., Lu, J. & Langmead, B. 2019. Improved metagenomic analysis with Kraken 2. Genome Biol, 20, 257.

24. World Health Organization (WHO) 2020. Director-General’s remarks at the media briefing on 2019-nCoV on 11 February 2020.

25. World Health Organization (WHO) 2023. Coronavirus disease (COVID-19) Weekly Epidemiological Update and Weekly Operational Update.

